# Machine Learning Prediction of Biomarkers from SNPs and of Disease Risk from Biomarkers in the UK Biobank

**DOI:** 10.1101/2021.04.01.21254711

**Authors:** Erik Widen, Timothy G. Raben, Louis Lello, Stephen D.H. Hsu

## Abstract

We use UK Biobank data to train predictors for 48 blood and urine markers such as HDL, LDL, lipoprotein A, glycated haemoglobin, … from SNP genotype. For example, our predictor correlates ∼ 0.76 with lipoprotein A level, which is highly heritable and an independent risk factor for heart disease. This may be the most accurate genomic prediction of a quantitative trait that has yet been produced (specifically, for European ancestry groups). We also train predictors of common disease risk using blood and urine biomarkers alone (no DNA information). Individuals who are at high risk (e.g., odds ratio of *>* 5x population average) can be identified for conditions such as coronary artery disease (AUC ∼ 0.75), diabetes (AUC ∼ 0.95), hypertension, liver and kidney problems, and cancer using biomarkers alone. Our atherosclerotic cardiovascular disease (ASCVD) predictor uses ∼ 10 biomarkers and performs in UKB evaluation as well as or better than the American College of Cardiology ASCVD Risk Estimator, which uses quite different inputs (age, diagnostic history, BMI, smoking status, statin usage, etc.). We compare polygenic risk scores (risk conditional on genotype: (risk score | SNPs)) for common diseases to the risk predictors which result from the concatenation of learned functions (risk score | biomarkers) and (biomarker | SNPs).

## 1. Introduction

Modern machine learning (ML) methods have opened the door to using high dimensional inputs to predict health outcomes and risk. This paper concerns the application of sparse linear ML to genetic and health information in order to make predictions that could be useful in a clinical setting. Recent work has highlighted that ML, especially polygenic predictors, have high potential impact in clinical settings [1–21]. The UK Biobank (UKB)[22] dataset includes single nucleotide polymorphisms (SNP) genotypes, medical diagnosis information, and extensive biomarker information (i.e., 48 quantitative outputs of blood and urine tests) for almost 500k individuals. In this article we describe ML investigations of the correlation structure between these three categories of data. As described in Figure 1, we train:

**Figure 1.**
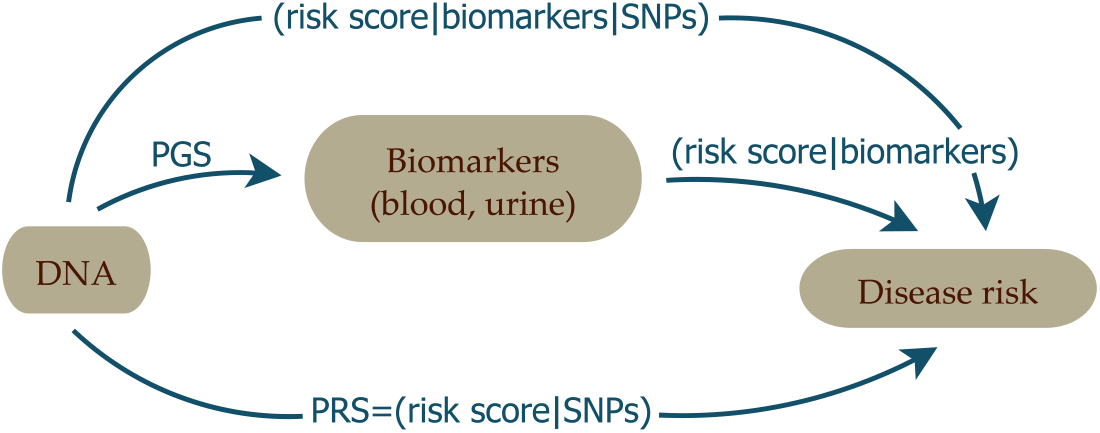
The four different types of predictors appearing in this paper.

1. Polygenic Score (PGS) predictors of the quantitative biomarker test results from *SNPs alone*. These functions predict biomarker level conditional on genotype: PGS = (biomarker | SNPs). For example, we predict measured lipoprotein A levels from SNPs, achieving a correlation of 0.76 between PGS and actual biomarker level. This may be the most accurate SNP prediction of a complex human trait yet accomplished.
2. Biomarker Risk Scores which predict risk of a specific disease condition *using only measured biomarkers as input*: (risk score | biomarkers). For example, our atherosclerotic cardiovascular disease (ASCVD) predictor uses ∼10 blood biomarkers to predict disease risk. We show that in UKB validation it predicts disease risk as well as or better than the American College of Cardiology ASCVD Risk Estimator [23,24], which uses quite different inputs such as age, diagnostic history, body mass index (BMI), smoking status, statin usage, etc. Liver and kidney problem risk prediction from biomarkers seems quite promising, based on our results. In total, we investigate predictions for ASCVD, coronary artery disease (CAD), diabetes type I & II, hypertension, very inclusive definitions of kidney and liver problems, and obesity.
3. Finally, by concatenating the predictors in 1 and 2 above, we build functions which map genotype (SNPs) to disease risk, with biomarkers as an intermediate step. We denote these concatenated predictors as: (risk score | biomarkers | SNPs). We emphasize that concatenation (i.e., *F*(*G*(*x*))) is *not* the same as training with both biomarkers and SNPs simultaneously used as features. The concatenated predictors *only* require SNPs as input, but use SNP predicted biomarker values as an *intermediate step* in calculation of the predicted disease risk. These functions can be compared to standard Polygenic Risk Scores (PRS) computed directly from SNPs, using disease case status as the training phenotype: PRS = (risk score | SNPs). For example, the concatenated function which maps SNPs*→* biomarkers*→* type 2 diabetes risk performs roughly as well as the PRS for type 2 diabetes (Area Under the Receiver operator characteristic Curve, AUC, ∼ 0.64).

From our investigations, we conclude that many biomarker levels are not just substantially heritable, but can be predicted with some accuracy from SNPs. This is true despite the fact that levels fluctuate from day to day for a specific individual! We also conclude that disease risk prediction from biomarkers alone, via (risk score | biomarkers), is potentially very powerful, and indeed complementary to existing methods for risk estimation. For example, we show below that existing ASCVD risk predictors use different and complementary information to the biomarkers used in our ASCVD (risk score | biomarkers). Our results suggest that combining this complementary information can lead to stronger prediction and perhaps new insights into heart disease. Significant analyses of the costs and benefits of additional inputs have been performed for the existing ASCVD predictor, which is in clinical use (e.g. [23,24]), including some of the features in our predictor. Our comparison is limited to risk predictor performance and in the UKB cohort only.

We validate all predictors using sibling data: most of the power to differentiate between siblings (either in quantitative trait values or disease risk) persists despite similarity in childhood environments. We also test the fall off in power in distant ancestries (relative to the European training population). The decline for SNP based predictors varies as expected with genetic distance, whereas biomarker prediction does not display this pattern.

Throughout this paper, we refer to the different biomarkers according to the abbreviations listed in Table 1.

**Table 1.**
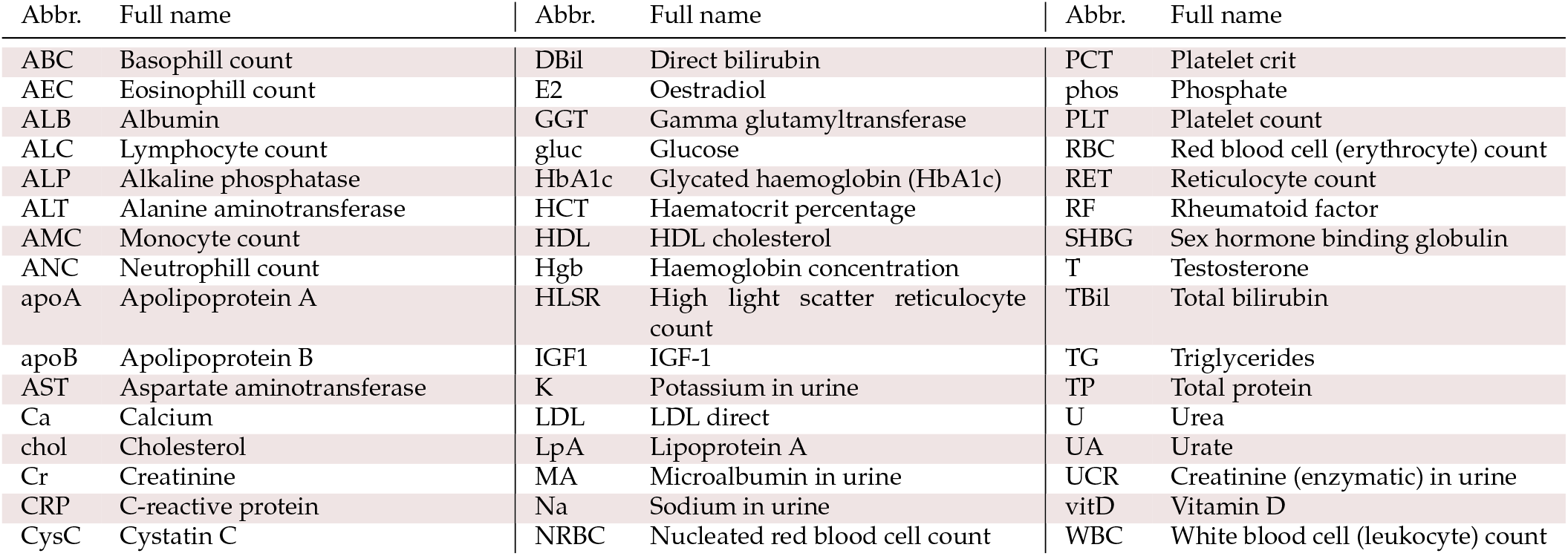
List of all studied blood and urine markers with abbreviations.

## 2. Materials and Methods

- Subject data All research in this paper uses data exclusively from the 2018 UKB release [22,25] and updates (see Supplementary Information for more details). All statements about sex or ancestry refer to the self-reported data within this dataset [26]. There is of course a complicated genetic substructure within each one of these subgroups [27–40], however, it has been repeatedly demonstrated that self-reporting provides sufficiently good data for training purposes [41–45]^1^. We refer to the self-reported ancestries labeled white, Asian, Chinese, and black in UKB as European, South-Asian, East-Asian and African, in accordance with the guidelines in [46]. It has been repeatedly confirmed that the power of polygenic predictors is dependent on both the training and testing ancestries, and that generally the power of the prediction falls off as a function of genetic distance [47–50]. All individuals with self-reported admixture were excluded from this study.
- Phenotype data The phenotypes included in the paper include self-reported UKB statuses, standard (ICD9, ICD10, OPCS3, OPCS4) codes, diagnosed conditions, thresholds, and combinations of all the previous items. Full details of how each phenotype is defined is given in the Supplementary Information.

### 2.1. Predicting Biomarkers from SNPs

This work primarily focuses on LASSO [51], or compressed sensing [52–55]. LASSO was chosen because it has been repeatedly shown that sparse, linear methods are among the most successful in genetic prediction over a wide variety of traits[11,41,43]. Additionally, sparsity makes application and analysis of the predictors much more computationally efficient. As genetic predictors move into clinical settings, it will undoubtedly be the case that optimal prediction algorithms will vary depending on phenotype and training data, but LASSO currently serves as an excellent jack-of-all-trades. We used LASSO to predict the 48 types of biomarkers listed in Table 1 from SNP data, and denote these type of predictors as (biomarker | SNPs).

- Data and pre-processing UKB contains data from repeated visits and for samples with more than one measurement of a certain biomarker the average value was taken. These raw measurements were z-scored for men and women separately and consecutively age corrected by subtracting a linear regression on age-biomarker data obtained from averaging the biomarker value for all samples born the same year (biomarker-age plots are contained in Supplementary Information). The parameters for the pre-processing were determined from training sets with about 340k samples of European ancestry. Evaluation sets of about 20-40k European siblings and all non-European individuals were withheld entirely from training but pre-processed with the same parameters. The UKB genotypic data were quality controlled by excluding all SNPs with less than 3% call success rate and also those with a minor allele frequency (MAF) 0.001. All individuals with less than 3% successfully called SNPs were also excluded and, again, any individual with self-reported mixed ancestry was excluded from this study entirely. Furthermore, only autosomal genetic information was used, including SNPs located on chromosomes 1-22 only.

- Predictor training Five LASSO predictors were trained on each biomarker using cross-validation, randomly drawing 1000 samples from the training set as validation set for each fold. The latter were used to choose optimal values of the regularization parameter *λ* (see Supplementary Information). The top performing predictor — as measured by correlation in the European corresponding validation set — from each fold was retained providing some statistics for the uncertainty estimates in the results. More details can be found in the Supplementary Informatio.

- Evaluation Each predictor for each biomarker was evaluated on its corresponding evaluation set consisting of ∼ 20-40k samples of European ancestry. To test the performance dependence on ancestry we also applied the predictors to the 9k of South-Asian, 1500 of East-Asian, and 7k of African ancestry. In section 3.2.2, we report the correlation between the PGS and the phenotypes as the performance metric for these continuous traits. Since environmental background, such as life style and diet, and indirect genetic effects have impacts on most of the biomarkers, we conducted a sibling evaluation. Siblings generally have more similar backgrounds than randomly chosen pairs, and are also more genetically similar than unrelated individuals. Retained predictive power among siblings is hence a strong indication of direct genetic effects. Moreover, the amount of lost power as compared to the general population can give some idea of the magnitude of environmental effects, e.g., from childhood environment. (There can also be genetic nurture [56–60] effects that are not analyzed here.) Childhood environments are more similar among siblings than between unrelated individuals, and this comparison gives an indication of whether the instantaneous biomarker measurements in adulthood are sensitive to the effects of childhood environment. To this end, we constructed both random pairs and pairs of genetic siblings within the evaluation set of European ancestry. For each pair, we calculated the difference in phenotype Δ_phen_ and the difference in PGS Δ_PGS_ and compared the correlations between these quantities corr(Δ_phen_, Δ_PGS_) within random and sibling pairs, respectively.

- Genetic architecture One can define the variance accounted for by each SNP *i* in a predictor according to

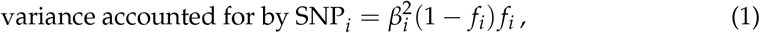

where *f*_*i*_ is the MAF of SNP *i*. This is described in greater detail in the Supplementary Information and in [42]. We use this alongside Manhattan-plots of the effect sizes *β* in the results (section 3.1.1) to display the genetic architectures of the top 3 performing (biomarker SNPs) predictors. Analogous plots for the rest of the (biomarker SNPs) predictors are contained in the Supplementary Information.

### 2.2. Methods for disease prediction

We used two approaches to investigate whether biomarkers can be used to predict disease risk, analogous to how blood tests are used clinically.

Approach 1: We trained predictors to predict case/control status directly from phenotypes, i.e., using the direct biomarker measurements as features. We denote this type of predictor (risk score | biomarkers), or biomarker risk score.
Approach 2: Second, we applied these already trained predictors to the *predicted* phenotypes, i.e., using the biomarker PGS output from the SNP-based predictors in section 2.1 as input. As such, we obtain disease risk scores using only SNP data as input. We denote these concatenated predictors (risk score | biomarkers | SNPs).

We evaluated this strategy on eight different condition definitions and we present the details for the two approaches separately. This is done both to display the performance dependence on the two approaches and since the prediction from biomarkers in approach 1 are very interesting in their own right.

#### 2.2.1. Approach 1: Predicting case status from biomarkers

- Condition definitions Based on the available UKB data, we defined conditions for CAD, cancer, diabetes type 1, diabetes type 2, hypertension, kidney problem, liver problem, and obesity. The detailed definitions for each one of these are to be found in the Supplementary Information. In general, we chose the definitions to be inclusive; kidney (liver) problem for example contains almost all kidney (liver) related problems that are reported in UKB, whereas cancer refers to any type of cancer. Obesity was defined as a BMI over 30. The effects of changing definitions are further discussed in section 4.
- Predictor training We used 45 out of the 48 biomarkers as input features, dropping E2, MA, and RF due to few available measurements, and taking the first available measurement for each sample. The raw data was pre-processed by sex specific z-scoring and then age correcting by subtracting a linear regression. Using LASSO, we then trained 5 predictors on the case/control status, choosing optimal *λ* by five-fold cross-validation. The training was done separately for men (*N* = 106, 656) and women (*N* = 86, 193) and on European ancestry only.
- Evaluation As was done for the (biomarker SNPs) in section 2.1, about 40k siblings of European ancestry and all non-European individuals were kept separate from all training and were used as evaluation set. We measured the predictor performance by AUC and by odds ratio plots. Additionally, we conducted sibling tests for the (risk score | biomarkers) predictors to test for environmental effects: we applied the predictors to pairs of siblings with precisely one case and one control and report the fraction of correctly called affected sibling, juxtaposed with the same results for random pairs of one case and one control. It should be emphasized here that we did not take date of onset into account in this study: disease status was considered on a “life span” (as far as UKB covers) basis such that cases could have onsets both prior to and after the time of the biomarker measurement. Prediction in this sense means what can we predict about current or future case status only knowing a set of momentary biomarker values. Temporal prediction tests (i.e., prospective prediction) are deferred to later work.

#### 2.2.2. Approach 2: Predicting case status from PGS of biomarkers

To form predictors taking SNP data as input, we concatenated the PGS predictors from section 2.1 with the biomarker predictors from approach 1, what we call (risk score | biomarkers | SNPs). The disease predictors (risk score | biomarkers) were taken as is from Approach 1 and applied to the z-scored PGS output of the predictors in section 2.1. No further training was done and the performance was evaluated as for and compared with the (risk score | biomarkers) predictors.

### 2.3. Comparison with ASCVD Risk Estimator

The ASCVD Risk Estimator [24] is a widely used tool to aid clinicians in risk estimations of and preventative care against atherosclerotic cardiovascular disease. We used this well-established resource to benchmark the approach of (risk score | biomarkers) predictors by training a predictor on this condition specifically. ASCVD aggregates several sub-diagnoses and exists in different versions. Hard ASCVD includes acute coronary syndromes, death by coronary heart disease, a history of myocardial infarction, and fatal and non-fatal stroke. A more general (extended) ASCVD definition additionally includes stable or unstable angina, coronary or other arterial revascularization, transient ischemic attack, and peripheral arterial disease presumed to be of atherosclerotic origin. We used a UKB specific extended definition, detailed in the Supplementary Information. The ASCVD Risk Estimator requires the input: age, sex, race, systolic and diastolic blood pressure, total cholesterol, HDL, LDL, history of diabetes, smoking status, time since quit smoking (if applicable), whether on hypertension treatment, whether on a statin, and whether on aspirin. It can also use previous data for follow-ups but we restricted our analysis to “first visit patients” only. All of these data fields can be found in some form in the UKB (the exact field choices are listed in the Supplementary Information).

The outputs of the ASCVD Risk Estimator are (up to) three risk estimates: 10 year risk, lifetime risk, and optimal risk, all given as a percentage. Since our UKB data only cover approximately 10 years from the first biomarker measurement, we exclusively used the 10 year risk output. We applied the underlying function of the ASCVD Risk Estimator to the corresponding data in UKB and obtained a 10 year risk estimate for 358,650 individuals for whom we also had an ASCVD case/control status. Strictly speaking, the ASCVD Risk Estimator was developed for North American cohorts and and based on hard ASCVD but, as seen in section 3.3, performed very well also in the cohorts of the UKB using the extended definition. Note, however, the current comparison is not intended as a rigorous test for deployment.

We then trained a (risk score | biomarkers) predictor on case/control status, analogously to approach 1 in section 2.2, but using ordinary linear regression on the z-scored biomarker measurements. This outputs a risk *score* which we mapped to absolute risk *estimates in percentages* as follows. The risk scores obtained from applying the predictor on the training data were binned and, within each bin, the disease prevalence was calculated from the case/control statuses as an estimated risk for samples with the corresponding risk scores. This discrete mapping was then made continuous using rolling averages and linear interpolation. For details see Supplementary Information.

#### 2.3.1. Combination of predictor from biomarkers and the ASCVD Risk Estimator

In the results section 3.3, we show that the ASCVD (risk score | biomarkers) predictor and the ASCVD Risk Estimator are making complementary predictions. We therefore also tested a combination of them. We made a linear regression on all the input features from the two predictors combined (48 continuous and 8 discrete variables), z-scoring the discrete variables from the ASCVD Risk Estimator input so that everything was on the same scale. In addition, we made a second regression also including the *output* of the ASCVD Risk Estimator to capture the non-linearities within that function. These regressions were made and evaluated on the same training and evaluation sets as for the (risk score | biomarkers) predictors.

## 3. Results

### 3.1. Predicting Biomarkers from SNPs

The performance of the (biomarker | SNPs) predictors ranges from the highest phenotype-PGS correlation for a polygenic predictor we are aware of to no predictive power whatsoever. We present the results in order of correlation within European ancestry in Figure 2. The best performing predictor is for lipoprotein A at a correlation of ∼0.76. This is not too surprising as lipoprotein A levels are well-known to be highly heritable [61–64], related to the LPA gene and other loci [65–74], and thus do not greatly vary by life style or environment ^2^. Yet, it is a striking example of predictive power. After lipoprotein A, we find correlations almost evenly distributed within the correlation range 0.1-0.59 and a group of 7 almost uncorrelated biomarkers at the bottom. In the same Figure 2, we have included the performance within the non-European ancestries. Being trained on European ancestry only, the predictors suffer the now familiar [49,50] fall-off pattern according to genetic distance, with performance generally being successively worse for South-Asian, East-Asian, and African ancestries.

**Figure 2.**
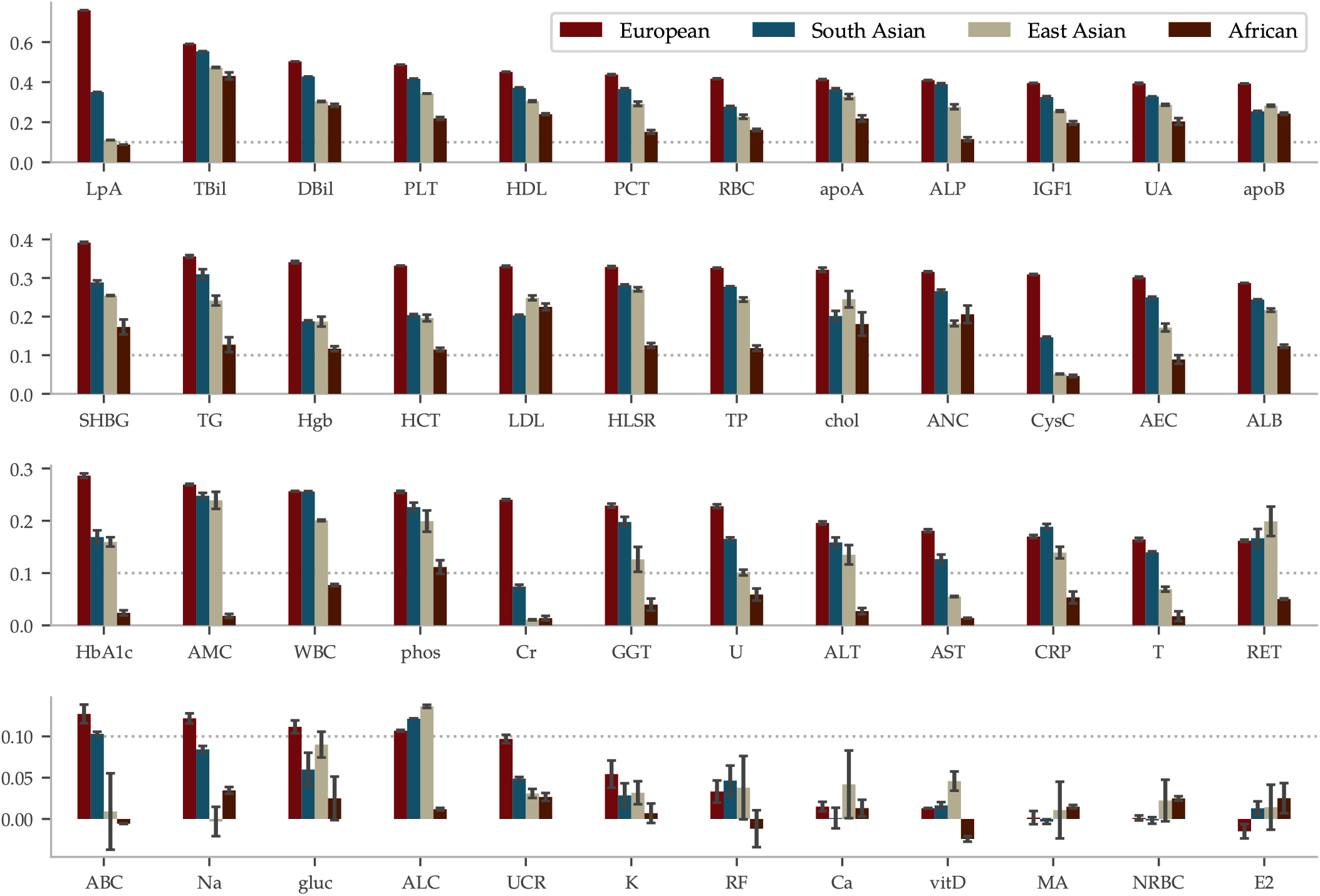
Correlations between PGS and phenotype vary from very strong to effectively zero, depending on the biomarker, and fall off with genetic distance from the training population. The mean of the PGS-phenotype correlation for evaluation sets are listed for all 48 biomarkers, ordered according to the results within Europeans — the ancestry for the training population. The error bars represent *±* the standard deviation for 5 different predictors trained on slightly different training sets. The dotted line is there to aid graphical comparisons across the rows. The LASSO predictor of lipoprotein A achieves a correlation of 0.759 within European ancestry, which is the highest correlation for a polygenic trait we are aware of. The correlation fall-off for the other ancestries generally follows the order European *>* South Asian *>* East Asian *>* African. Note that the sample sizes for these ancestries are much smaller.

The results from the sibling comparison can be seen in Figure 3. On average, there is a ∼26% drop in correlation when comparing differences within random pairs and differences within sibling pairs. The figure also shows that siblings that are separated by more than 0.5, 1.0, and 1.5 times the standard deviation in phenotype are predicted with increased correlation. The sibling comparisons for the other biomarkers can be found in the Supplementary Information.

**Figure 3.**
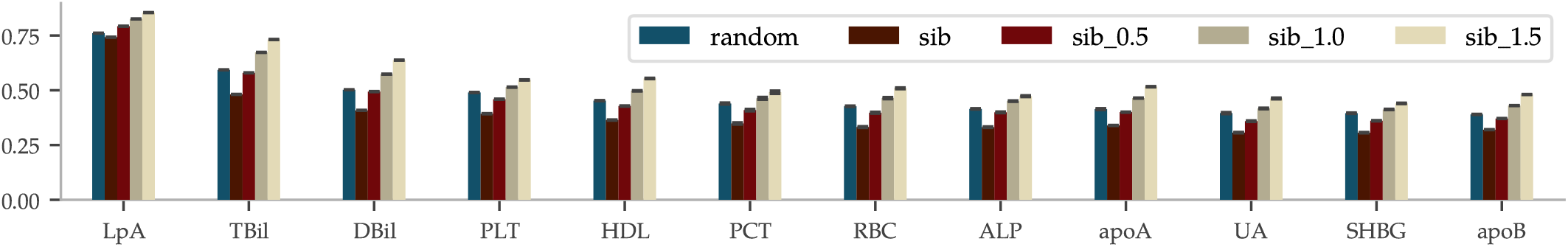
Sibling comparisons of correlation between difference in phenotype and difference in PGS, i.e., corr(Δ_phen_, Δ_PGS_), show that most of the correlation is retained also for pairs that share similar environmental backgrounds. UKBs 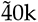 siblings of European ancestry were paired either randomly or as genetic siblings and were used as test set. The correlations between the pairs’ differences in phenotype and their differences in PGS was then calculated for each biomarker, ordered above from strongest to weakest correlation. The error bars indicate *±* the standard deviations for 5 predictors trained on slightly different training sets. The additional three bars 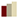 labeled sib 0.5, sib 1.0 and sib 1.5, are the results when restricting to siblings with phenotype differences larger than 0.5, 1 and 1.5 standard deviations, respectively. Two siblings are likely to have more similar environmental backgrounds than random pairs, affecting the similarity of late-life biomarker measurements independently from (direct) genetic effects. This could explain the decreased correlation for siblings as compared to random pairs. Yet, the remaining correlations are strong evidence that the predictors capture some direct genetic effects on the biomarkers. The comprehensive figure for all biomarkers can be found in the Supplementary Information.

#### 3.1.1. Genetic Architecture

Polygenic predictors have shown to usually use information spread over the entire genome, even when enforcing sparsity [11,41,42,45]. In Figure 4, we illustrate the genetic architectures behind three of the top performing (biomarker | SNPs) predictors with Manhattan plots of the effect sizes *β* and the variance accounted for in eq. (1), accumulated across chromosomes 1-22 (the Supplementary Information contains figures for all biomarkers). It shows that both biomarkers with a few very strong loci and biomarkers with an evenly distributed dependence can be predicted well. Let us make a few remarks on the top 5 performing predictors (see Supplementary Information for the direct bilirubin and platelet count plots):

**Figure 4.**
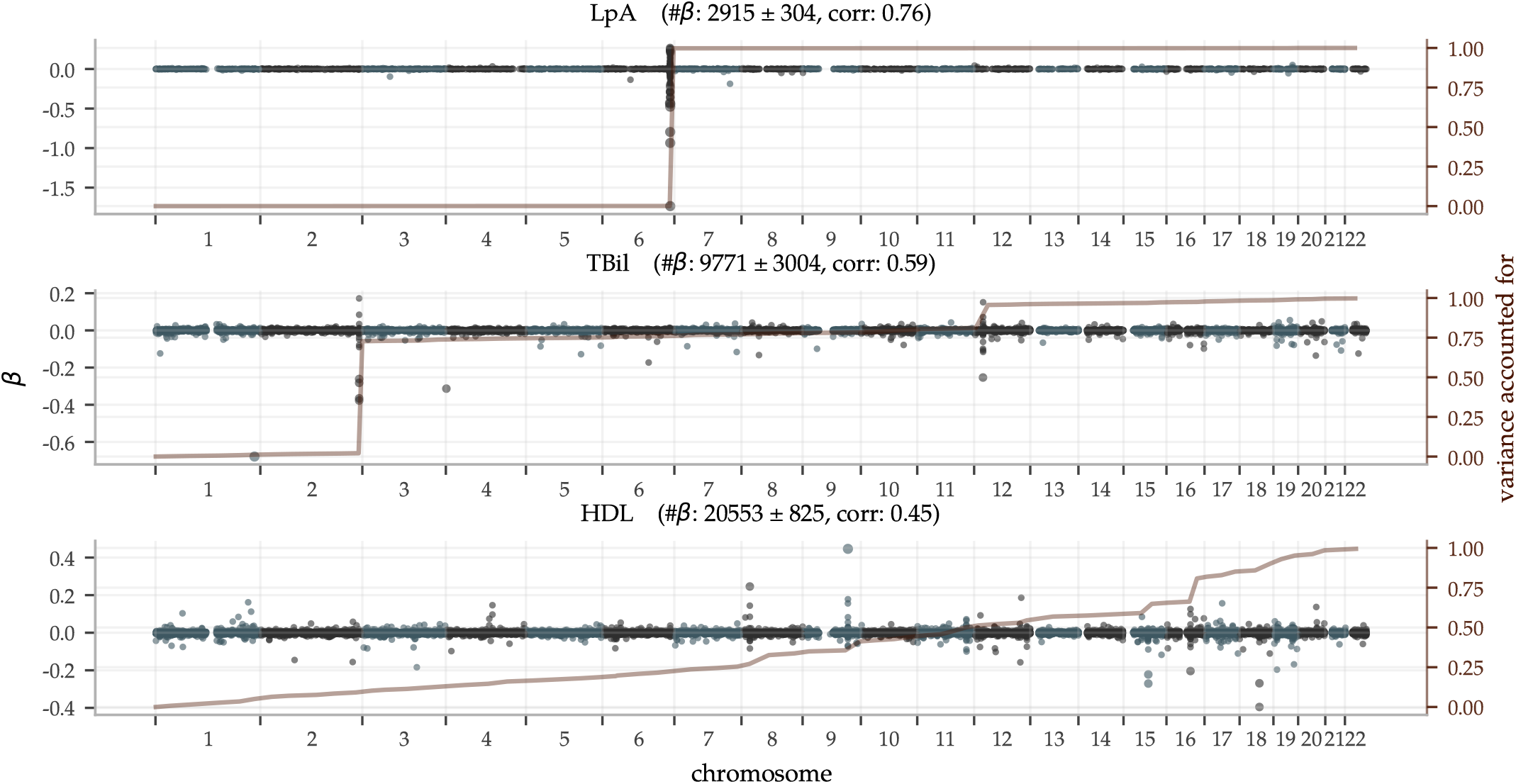
Manhattan plots of LASSO *β* — superimposed with the aggregate single snp variance accounted for — show both highly localized as well as widely polygenic architectures. The predictor for Lipoprotein A is almost entirely determined by the well-known gene LPA in chromosome 6; the top 50 SNPs in this region account for ∼ 95% of the aggregate single SNP variance. In contrast, HDL has an almost uniform distribution of the variance accounted for across all the 22 autosomal chromosomes, despite some loci with high magnitude *β*-coefficients. (The difference being due to the MAF in equation (1).) The most significant genetic loci are discussed further in the main text. The plot titles include the achieved PGS-phenotype correlation and mean number of non-zero *β ±* the standard deviation for the 5 predictors trained on each trait.

- The lipoprotein A predictor is as expected totally dominated by the single locus on chromosome 6, the gene carrying its name LPA.
- The total bilirubin predictor is very similar to the one for direct bilirubin. GWASes have implicated many variants on all but chromosome 15 (according to a GWAS Catalog[76] trait search) but most have a very minor impact on our predictor. For example, [77] reported a locus on chromosome 19 but although there are groups of moderately large *β* in this region, the entire chromosome 19 does not account for more than ∼ 1% of total variance in our predictors.
- GWASes for direct bilirubin in the literature [77,78] are generally dominated by variants in gene UGT1A on chromosome 2. The LASSO predictors pick these up too. In addition, there is another ∼ 17% variance accounted for by the locus at chromosome 12, also known[78]. Chromosomes 6 and 19 account for ∼ 1% variance each and have no generally listed loci. The *β*_*i*_ with the largest magnitude corresponds to SNP rs908327 on chromosome 1. It has SNPs in linkage disequilibrium (LD) that have been linked to triglycerides[79] but not directly to bilirubin, to our knowledge. It has a very small MAF, however, and does not account for much variance.
- The predictor for platelet count is very polygenic with the variance accounted for almost evenly distributed across all 22 chromosomes. Chromosome 12 provides a small deviation from this pattern, accounting for ∼14% of the variance, partly due to a locus near one end.
- The predictor for HDL is also highly polygenic. Previous GWASes have recorded loci at all but chromosome 13, which has no large magnitude *β*_*i*_ but still accounts for ∼ 1% of the total variance.

### 3.2. Predicting Disease Risk

The results for the disease risk predictors are divided into sections corresponding to the (risk score | biomarkers) from approach 1 and (risk score | biomarkers | SNPs) from approach 2, respectively.

#### 3.2.1. Predicting case status from biomarkers

The performance of the (risk score | biomarkers) predictors was evaluated and are reported as AUCs and odds ratio plots in Figure 5. With training optimized for European ancestry, we regard the results for this ancestry as the main results and provide the performance in other ancestries for reference. The results vary with the condition. Within European ancestry, they range from an AUC of .53 (.60) for cancer for women (men) up to ∼ .95 for diabetes type 1 (both sexes). As a comparison, we report below on an ASCVD predictor with an AUC of ∼ .76 which performs risk prediction as well as or better than the American College of Cardiology ASCVD Risk Estimator. We discuss this in detail below in section 4. The odds ratio plots show a wide range of results that also vary with condition. Figure 5 separates conditions into groups based on the odds ratios of the high risk outliers. The strength of the diabetes predictors is probably due to their use of blood biomarkers (e.g. HbA1c) which are standard diagnostic indicators for diabetes. That this standard diagnostic indicator is so highly ranked lends confidence to the results of the general methodology.

**Figure 5.**
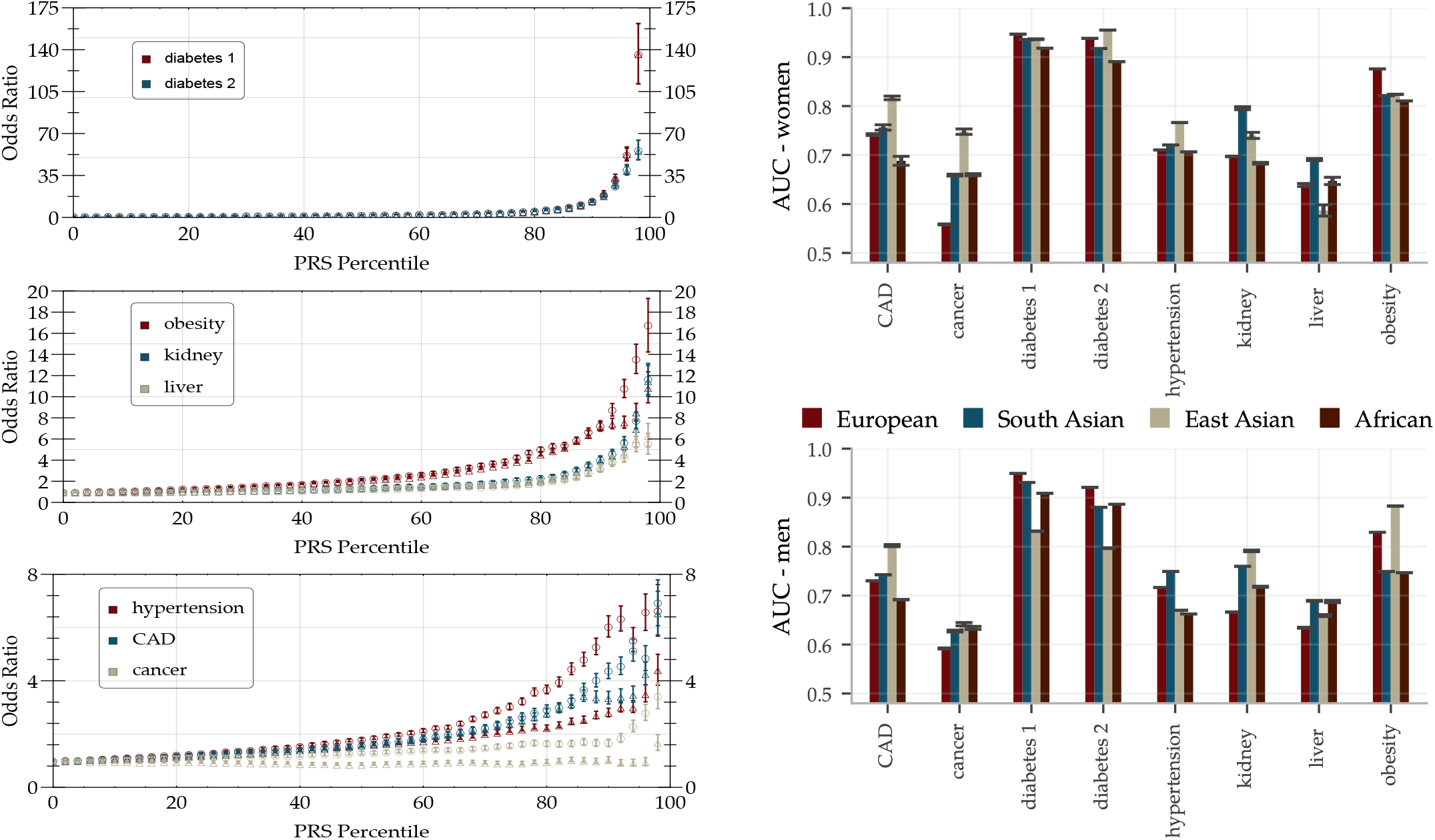
The predictive power of (risk score | biomarkers) can single out high risk individuals with over 10x odds ratio for many traits, and AUCs *>* 0.7 for most traits including tests across ancestry. **Left:** inclusive odds ratio (OR) plots for diabetes type 1/2, obesity, kidney problem, liver problem, hypertension, CAD, and any cancer trained and validated on the European population. Horizontal axis indicates individuals at that percentile *and above* in PRS. Marker is for predictors trained and validated on men and marker Δ for predictors trained and validated on women. Error bars represent the standard error of the mean value with a contribution coming from computing the OR and a contribution from including 5 predictors. **Right:** AUCs for (risk score | biomarkers) predictors separately trained on men and women. All predictors are trained on the European population and then validated on European, South Asian, East Asian, and African populations. The error bars indicate the standard deviations for 5 different predictors and do not reflect the significant uncertainties arising from limited available statistics (sample sizes are listed in Supplementary Information).

There are some differences in performance for men and women, most notably in cancer (possibly due to sex specific cancer variants). The differences are condition specific and viewed across all conditions the performance is similar. We delay a more detailed analysis of these differences to future study. The reported performance variations across the different ancestries are notably smaller and show less of a consistent pattern than what is the usual case for prediction from genetic information; this is expected since predicting from biomarkers stays on a higher biological level and does not involve issues such as LD patterns and tag SNPs etc. Note, however, that these results are limited by the available statistics, see Supplementary Information for the case/control numbers for each ancestry.

In Figure 6, we also include two examples of the LASSO coefficients for CAD and type 2 diabetes. For CAD, we find mostly well-known biomarkers with the highest weight, such as LDL, apolipoprotein B, total cholesterol and HDL. However, for women cystatin C appears at fourth place, which to our knowledge is not often used in this context. Cystatin C also is the fifth most influential biomarker in the diabetes type 2 predictor for both sexes, while these predictors are dominated by the standard biomarker glycated haemoglobin. In fact, cystatin C is among the more important biomarkers for most of our predictors. Coefficients for all conditions are listed in the Supplementary Information.

**Figure 6.**
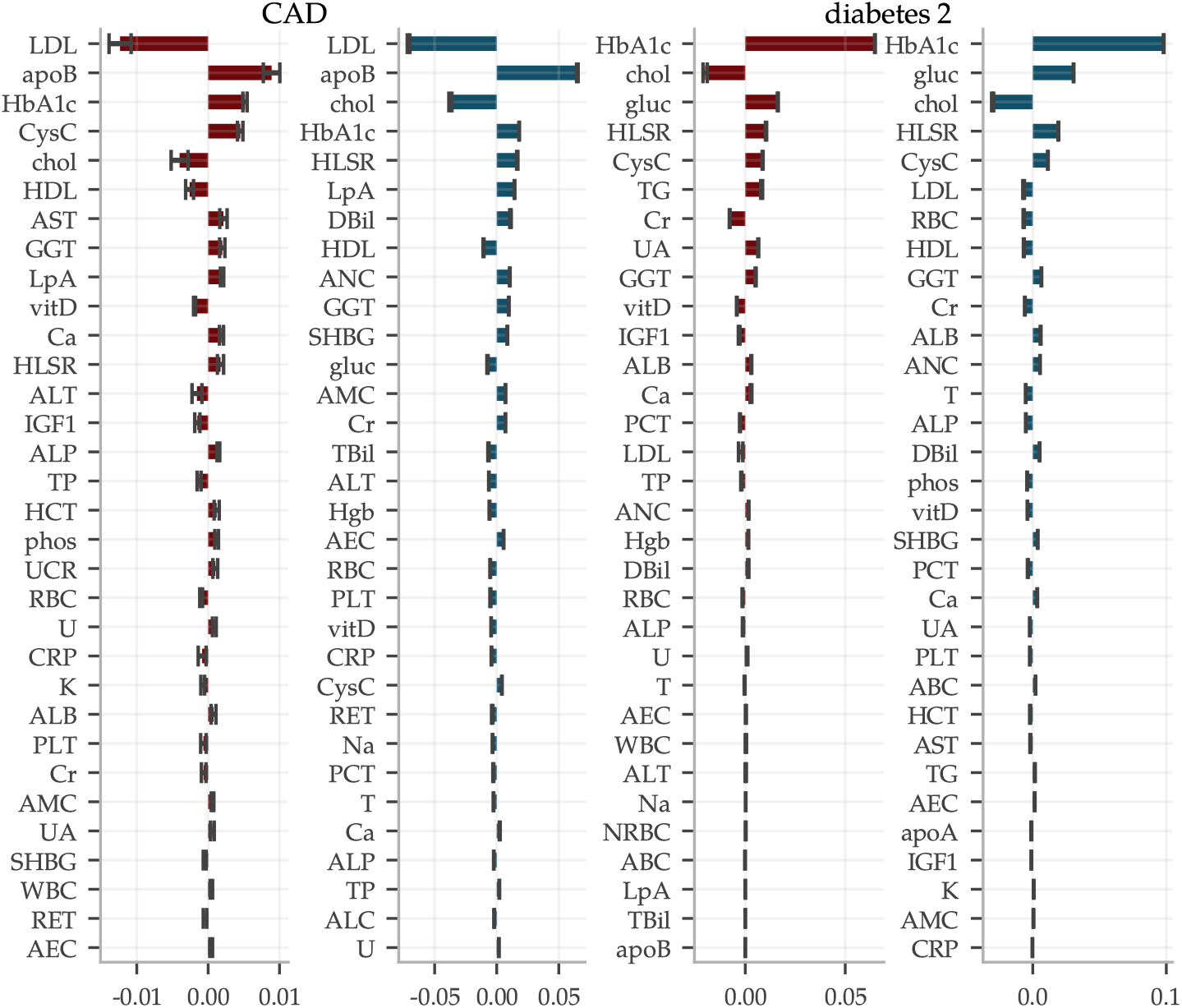
Predictors for phenotypes like CAD and type 2 diabetes from biomarkers are dominated by a top few inputs. Relative weights of each biomarker within predictors for CAD and type 2 diabetes. 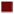 women and 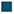 men while error bars indicate *±* standard deviations from the mean of five predictors. The most impactful biomarkers are very well-known but we highlight cystatin C as surprisingly frequent among the moderately strong coefficients. Corresponding plots for all condition predictors are shown in the Supplementary Information.

We investigated the presence of non-linear effects for (risk score | biomarkers) by extending the input features with all possible quadratic interactions among the seven most influential biomarkers for each condition. We saw no effect on the performance in either direction and conclude that the effects of the biomarkers on all the listed conditions appear to be linear to very good approximation.

#### 3.2.2 Predicting case status from PGS of biomarkers

The concatenated predictors (risk score | biomarkers | SNPs) suffer a significant drop in performance, as can be seen in Figure 7. The imprecise predictors (biomarker SNPs) introduce a lot of noise and, exacerbated further by the uncertainty in the (risk score | biomarkers) predictors, the concatenation does in general not lead to meaningful predictions. A notable exception are the diabetes predictors. The combination of reasonably correlated PGS for the most important biomarkers and the exceptionally high AUCs for these predictors lead to an average AUC of ∼ .63 for the type 2 diabetes (risk score | biomarkers | SNPs) predictor. This is comparable to what we have achieved in the past by training SNP-based LASSO directly on type 2 diabetes status[11]. Furthermore, the two different types of predictors (risk score | biomarkers | SNPs) and (risk score | SNPs) capture somewhat complementary information, as shown in Figure 8. The sum of the two types of risk scores reaches an AUC of ∼ .67. It is unclear why the use of biomarkers as an intermediate step adds additional information relative to training directly with SNPs as features and case status as the phenotype. We leave this as an interesting topic for future research.

**Figure 7.**
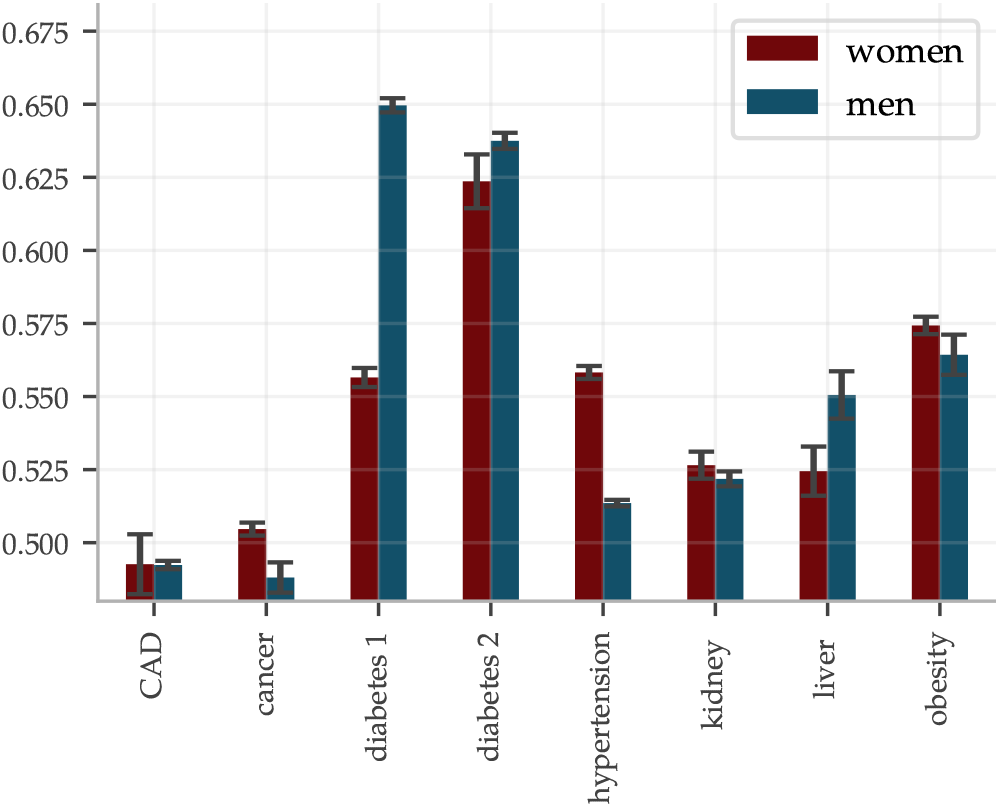
AUCs for (risk score | biomarkers | SNPs) predictors drop significantly as compared to (risk score | biomarkers) in Figure 5 and only the diabetes predictors reach par with other methods. The predictors were evaluated on 9016 (9607) white women (men) and the error bars indicate *±* the standard deviation for 5 different predictors.

**Figure 8.**
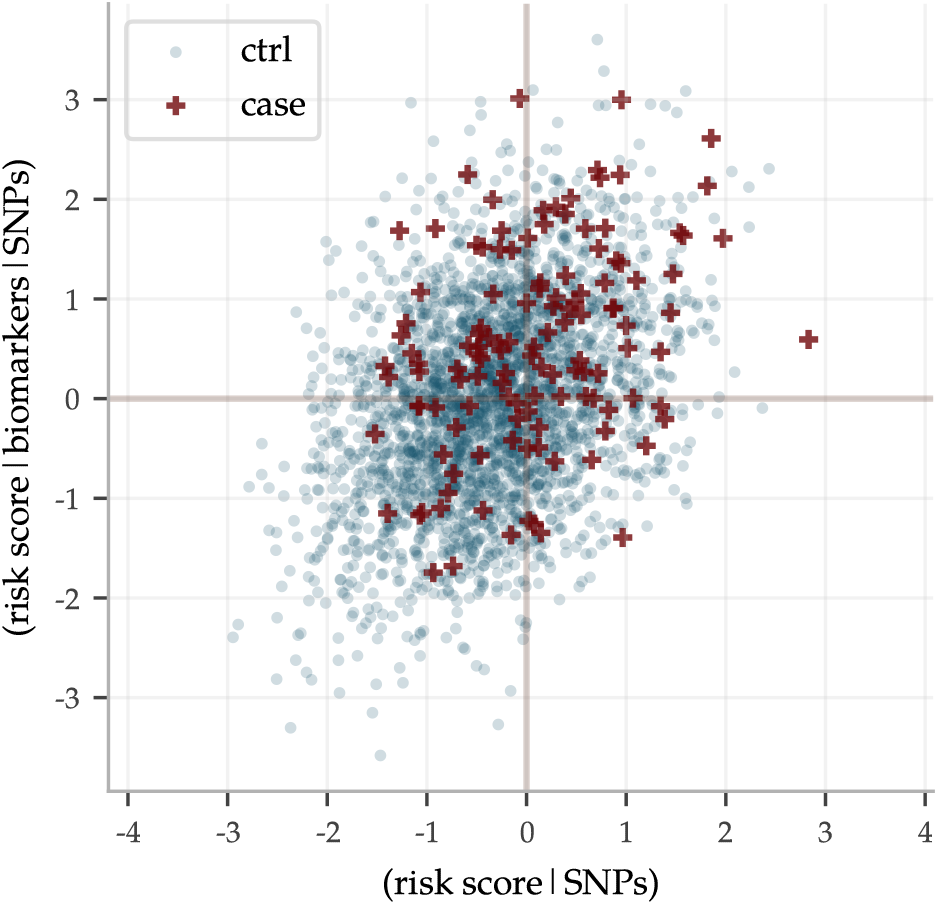
Risk scores predicted from SNPs (risk score | SNPs) and from PGS of biomarkers (risk score | biomarkers | SNPs) do not always agree, here exemplified by type 2 diabetes data for men. Both predictors predict case status directly from SNPs alone. Their outputs correlate ∼ 0.37 with a linear regression coefficient of ∼ 0.39. In the noise, they capture some complementary information: the sum of the risk scores achieves an AUC of ∼ 0.67 while the SNP and PGS based predictors individually achieve AUCs of ∼ 0.63 and ∼ 0.65, respectively.

The sibling evaluation of the disease risk predictors, described in section 2.2, is reported in Figure 9. The fraction of sibling pairs with one case and one control called correctly ranged from pure chance for cancer and liver problems, while reaching ~ 0.9 for diabetes type 1 and 2, using the (risk score | biomarkers) predictors. The accuracy dropped significantly for the (risk score | biomarkers | SNPs) predictors, as expected; no predictor of this type reached a correctly called fraction above 0.6.

**Figure 9.**
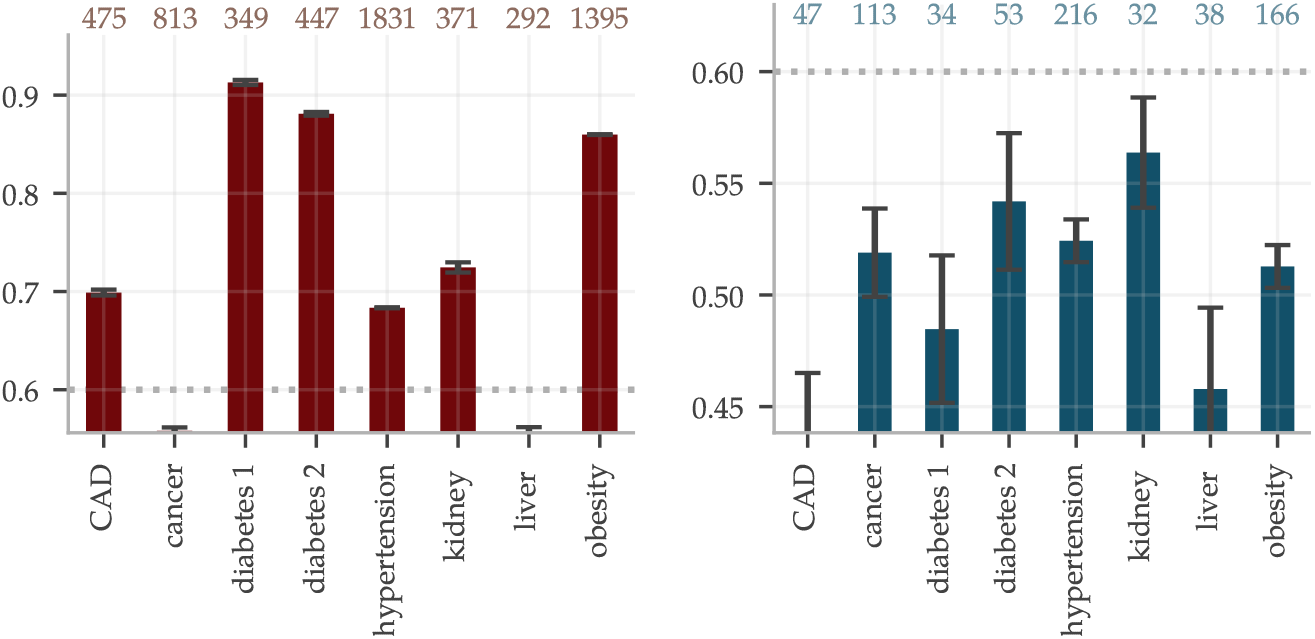
The fractions of sibling pairs with precisely one case and one control called correctly are generally high for 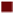 (risk score | biomarkers) but not much better than chance when predicting from genotypes using 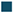 (risk score | biomarkers | SNPs). The pairs were considered correctly called if the PRS was higher for the affected sibling, without any restriction on the size of the separation. Number of included sibling pairs differed for the two types of predictors and are listed at the top. The error bars indicate *±* the standard deviation for five different predictors for (risk score | biomarkers) and for 5 *×* 5 concatenation combinations of predictors in the (risk score | biomarkers | SNPs).

### 3.3. Comparison with ASCVD Risk Estimator

To illustrate the performance of the (risk score | biomarkers) predictor for ASCVD and to compare it with the ASCVD Risk Estimator, we used the risk percentage output as described in section 2.3. The ASCVD Risk Estimator was built using American cohorts of separately European and African ancestry. Due to the similarities with the UKB population, we deemed it could be applied somewhat fairly to the entire UKB, whereas we used the withheld evaluation set of ∼ 40k of European ancestry for the (risk score | biomarkers) predictor. The result is shown in Figure 10, in which the predicted risks were binned and the actual disease prevalence within each bin was calculated, labeled “Actual risk”. Both predictors give very accurate risk estimates, with increasing uncertainty for individuals with high predicted risk. However, although they do assign correct risk estimates for bins taken as a whole, they do not always agree on who is at low versus high risk. The scatter plot in Figure 10 shows their individual distributions and occasional disagreements. Their partially complementary predictions are further highlighted in the risk heat map in Figure 10 and utilized below in a combined predictor.

**Figure 10.**
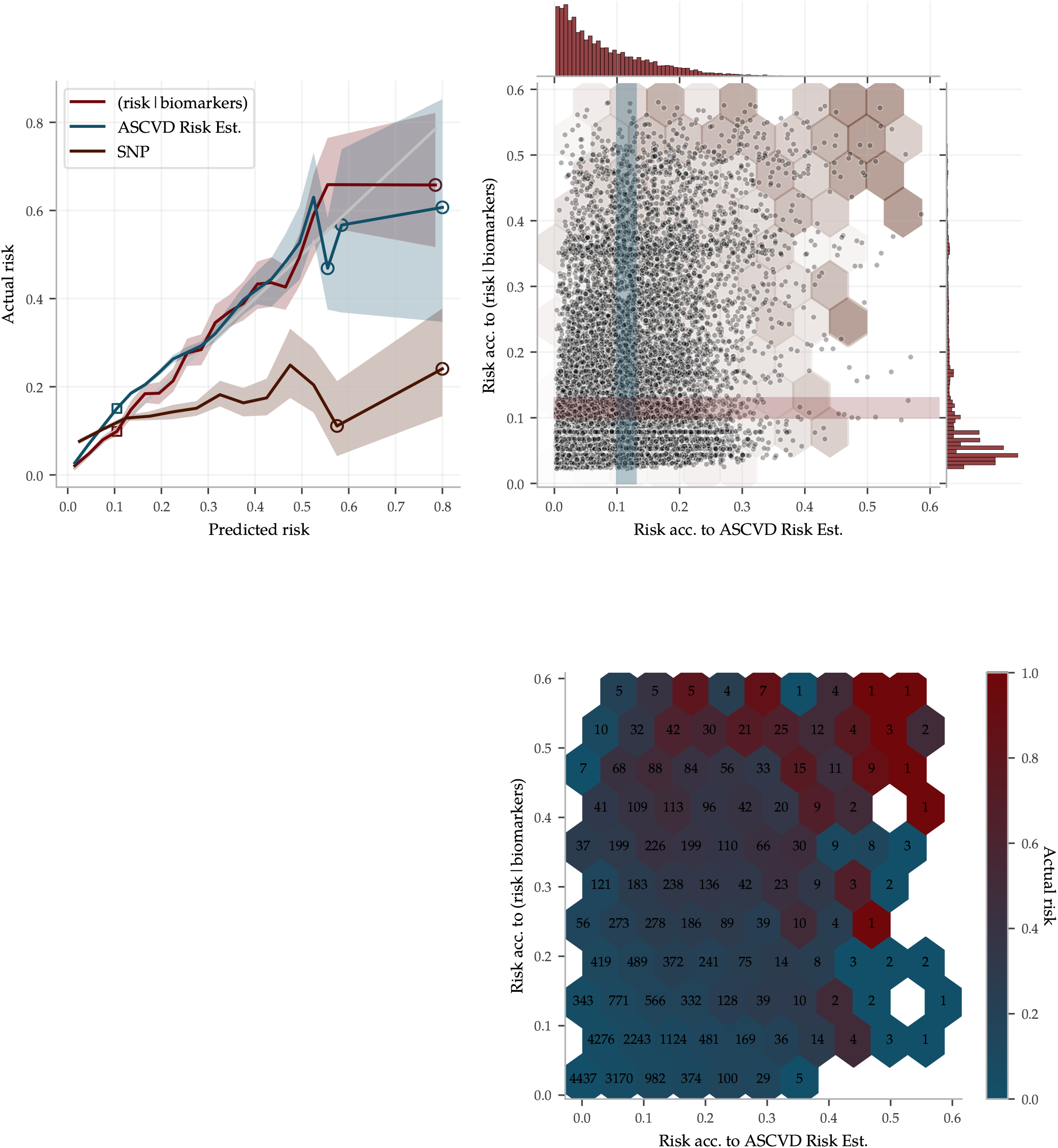
The ASCVD (risk score | biomarkers) and the ASCVD Risk Estimator both make accurate risk predictions but with partially complementary information. **Left:** Predicted risk by (risk score | biomarkers), the ASCVD Risk Estimator and a (risk score | SNPs) predictor were binned and compared to the actual disease prevalence within each bin. The gray 1:1 line indicates perfect prediction. Shaded regions are 95% confidence intervals obtained from 100 fold bootstrap estimates of the prevalence in each bin. The ASCVD Risk Estimator was applied to 340k UKB samples while the others were applied to an evaluation set of 28k samples, all of European ancestry. **Upper right** shows a scatter plot and distributions of the risk predicted by (risk score | biomarkers) versus the risk predicted by the ASCVD Risk Estimator for the 28k Europeans in the evaluation set. The (risk score | biomarkers) distribution has a longer tail of high predicted risk, providing the tighter confidence interval in this region. The left plot y-axis is the actual prevalence within the horizontal and vertical crosssections, as illustrated with the shaded bands corresponding to the hollow squares to the left. Notably, both predictors perform well despite the differences in assigned stratification. The hexagons are an overlay of the **lower right** heat map of actual risk within each bin (numbers are bin sizes). Both high risk edges have varying actual prevalence but with a very strong enrichment when the two predictors agree.

#### 3.3.1. Combination of predictor from biomarkers and the ASCVD Risk Estimator

Since the ASCVD Risk Estimator and the (risk score | biomarkers) predictor use different input and give complementary predictions, we combined them into a a very reliable risk predictor, superseding both the former. The risk estimates are compared with actual disease prevalence in Figure 12 for two versions of the combined predictor: *(1)* a linear regression on the biomarkers and all of the input going into the ASCVD Risk Estimator, and *(2)* a similar regression but also including the *output* of the ASCVD Risk Estimator. Their top coefficients are listed in the same figure.

**Figure 12.**
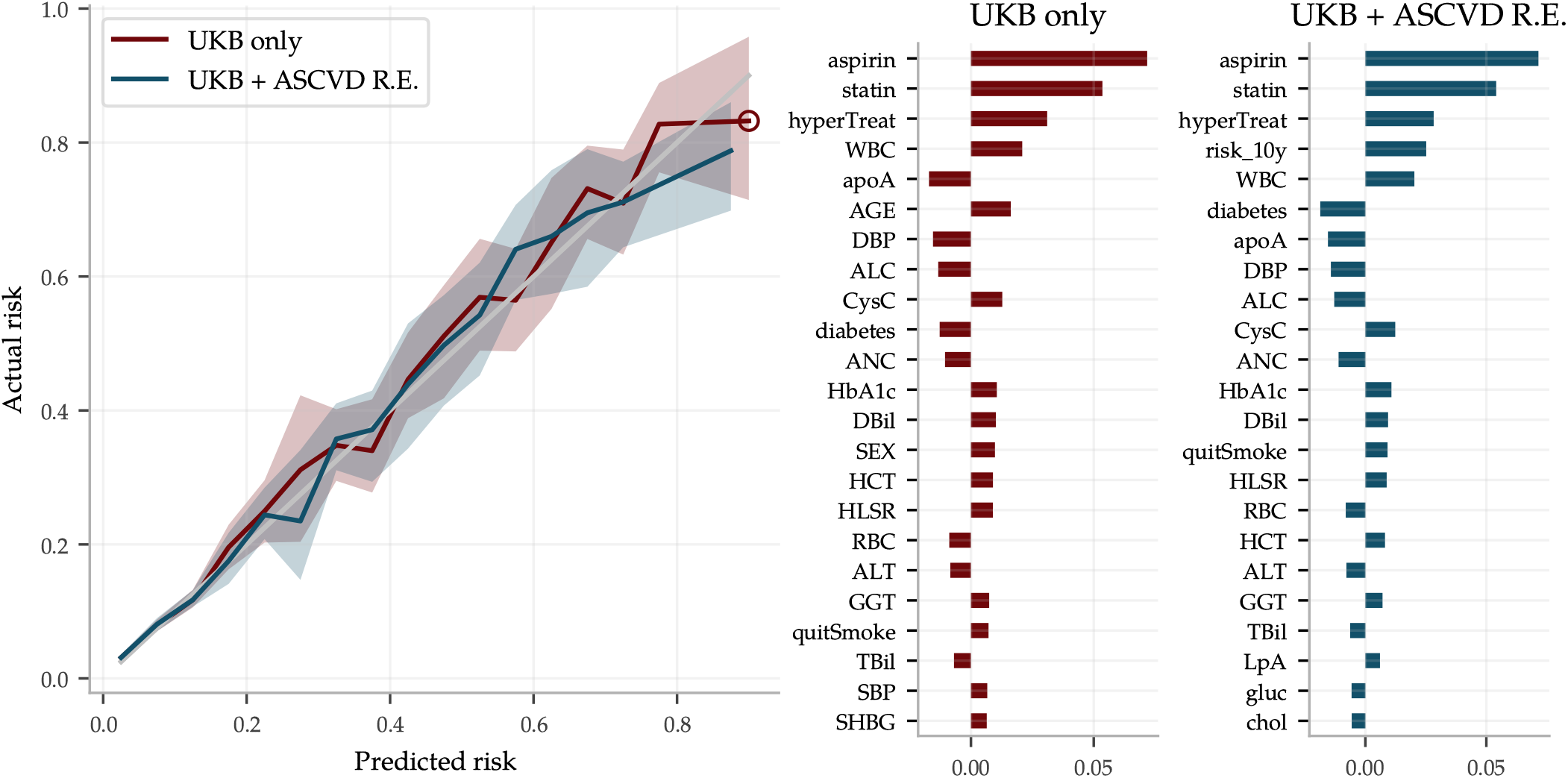
The risk prediction using both 45 biomarkers and all the ASCVD Risk Estimator input improves performance as compared to Figure 10, in particular for high risk individuals, and is very good all the way up to risk levels of 80%. The figure compares two predictors: 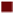 a combined ASCVD predictor using all 45 biomarkers plus all the input fields (age, sex, etc.) used by the ASCVD Risk Estimator, using UKB data only, and 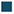 a predictor using the same input plus the ASCVD Risk Estimator *output*, labeled UKB + ASCVD R.E. The latter does not perform notably better, although the ASCVD Risk Estimator output “risk_10y” corresponds to the fourth strongest coefficient. Both perform better than both the (risk score | biomarkers) and ASCVD Risk Estimator individually, confirming their complementary nature shown in the heat map, Figure 10. The shaded areas in the left panel again indicate 95% confidence intervals obtained by 100 fold bootstrap calculations of the actual prevalence in each risk bin. Figures with all coefficients can be found in the Supplementary Information.

## 4. Discussion

UK Biobank data include about 500k individuals, for each of whom the following are recorded: SNP genotype, biomarker (blood, urine) test results, and case status for most common disease conditions. We have explored the pattern of correlations between these three distinct data types using machine learning.

We have shown that SNPs can be used to predict quantitative values of biomarkers by training new polygenic scores (PGS) for biomarker prediction. We note that the day to day fluctuation of these biomarker levels suppresses the quality of prediction. A more stable phenotype (e.g., average value of biomarker measured on multiple occasions) would probably be even better predicted from SNPs alone.

As is typical for current genomic predictors, we find predictive power falls off significantly with genetic distance from the (European) training population. This highlights the importance of increasing ancestry diversity in genetic data collection. As genetic predictors begin to find clinical applications, lack of diversity can exacerbate healthcare inequalities[48,80] (a larger list of associated ethical issues is highlighted in [44]).

We showed that biomarkers can be used as input to predict common disease risk. Some of these (risk score| biomarkers) predictors (e.g., ASCVD, diabetes) are very strong and may even surpass risk predictors in widespread clinical use. The combined predictor trained using both biomarkers and ASCVD Risk Estimator inputs clearly outperforms the latter in our comparison, at least for individuals at very high risk. It should be emphasized here that we did not perform the careful evidence review nor the statistical analysis that underlie the ASCVD Risk Estimator [23] and our comparison did not take into account the time of diagnosis. As such, our ASCVD predictor presented here is merely a comparative example and is not intended for clinical use in its current form. Yet, this naive approach performs remarkably well, utilizing the large statistical power of the UKB.

In the case of liver and kidney disease, we are not aware of other quantitative risk predictors that can be evaluated from biomarkers alone. Our results suggest that further research in this direction is warranted.

We note that (risk score | biomarkers) prediction quality does not exhibit the pattern of fall-off with genetic distance as previously found with genomic predictors^3^. For example, CAD and ASCVD predictors work well in all major ancestry groups despite using a European training sample. Further investigation is needed.

We studied concatenated predictor functions, which map SNPs to biomarkers to risk. In general, there were significant declines in performance. The magnitudes of these declines were perhaps expected for correlation chains of generic, high dimensional, vectors with similar pairwise correlations. Of the (risk score | biomarkers | SNPs) predictors, only the type 2 diabetes predictor performs well: AUC of ∼ .63. This is in fact comparable to what we have achieved in the past by training SNP-based LASSO directly on type 2 diabetes status. Furthermore, the two different types of predictors (risk score | biomarkers | SNPs) and (risk score |SNPs) capture somewhat complementary information, as shown in Figure 8. The sum of the two types of risk scores reaches an AUC of ∼ .67. It is unclear why the use of biomarkers as an intermediate step adds additional information relative to training directly with SNPs as features and case status as the phenotype. We leave this as an interesting topic for future research.

## Data Availability

The predictors described in the paper are available to other researchers
upon request.

## Author Contributions

Authors, listed alphabetically according to last name, contributed in the following ways: conceptualization, L.L., S.H. and E.W.; methodology, L.L., T.G.R., S.H., E.W.; software, L.L., T.G.R., and E.W.; validation, L.L., T.G.R., E.W.; formal analysis, L.L., T.G.R., E.W.; investigation, L.L., T.G.R., E.W.; resources, S.H.; data curation, L.L., T.G.R., and E.W.; writing — original draft preparation, L.L., T.G.R., S.H., and E.W.; writing — review and editing, L.L., T.G.R., S.H., E.W.; visualization, T.G.R. and E.W.; supervision, S.H.; project administration, S.H.; funding acquisition, S.H.; All authors have read and agreed to the published version of the manuscript.

## Data Availability Statement

The predictors described in the paper are available to other researchers upon request.

## Acknowledgments

Computational resources provided by the Michigan State University HighPerformance Computing Center. The authors thank Dr. Andrew Siskind (UC Irvine) and Dr. Pui-Yan Kwok (Academia Sinica UC San Francisco) for fruitful discussions and correspondence. The authors acknowledge acquisition of datasets via UK Biobank Main Application 15326.

## Footnotes

1 Genetic prediction in general depends on non-trivial factors including population substructure, size of training sets, algorithms (e.g. sparse vs non-sparse methods), heritability, environmental factors, and etc. Nonetheless, in many instances self-reported identity is sufficient for training.

2 Lipoprotein A has long been studied because of its association with CAD, atherosclerotic risk, liver problems, metabolism, and even cancer. Further discussion can be found in the review [75]

3 Previous GWAS and PGS studies generally see a fall off behavior, but there are occasional exceptions (e.g. [81]).

