## Supplementary Information for "Machine Learning Prediction of Biomarkers from SNPs and of Disease Risk from Biomarkers in the UK Biobank"

### Supplementary Information — Machine Learning Prediction of Blood and Urine Biomarkers from Genotype and of Disease Risk from Biomarkers in the UK Biobank

Erik Widen Timothy G. Raben Louis Lello Stephen D.H. Hsu

April 1, 2021

#### S0 Contents

**S1 Methods** **S1**
S1.1 LASSO . . . S1

**S2 Phenotype Descriptions** **S2**
S2.1 Phenotype distribution by sex for European ancestry . . . S2
S2.2 Phenotype dependence on age . . . S5

**S3 Phenotype Prediction** **S7**
S3.1 Calibration . . . S7
S3.2 PGS correlations . . . S7
S3.3 Sibling evaluation . . . S9
S3.4 Genetic Architectures . . . S10

**S4 Disease Prediction** **S17**
S4.1 Disease condition definitions . . . S17
S4.2 Input data to the ASCVD Risk Estimator . . . S19
S4.3 Risk conversion for ASCVD predictors . . . S19
S4.4 Coefficient sizes . . . S19
S4.5 AUCs and sample sizes . . . S23
S4.6 Risk score distributions . . . S25

#### S1 Methods

Here we expand upon the sparse prediction methods described in the main text. Things like how error bars were calculated, how we used python and cross checked things with julia, what type of computer cluster we ran on, etc.

##### S1.1 LASSO

Implementing LASSO amounts to finding the set of $p$ coefficients $\beta$ that minimizes the objective function

$$\mathcal{O} = \frac{1}{2}\|\hat{y} - X\beta\|^2_{L_2} + \lambda|\beta|_{L_1}\,, \tag{1}$$

where $X$ is the $(N \times p)$ genotype matrix with the number of minor alleles at each SNP position for each sample and $\lambda$ is a hyperparameter controlling the amount of regularization imposed on $\beta$. $\hat{y}$ is the $N$-vector of phenotype values for each sample which is either a raw phenotype, $y$, or adjusted via linear regression for age, sex, other covariates, or environmental factors (E), i.e. $\hat{y} = y - \mu - EW$, for regression coefficients $\mu$ and $W$. Applying the predictor to a genotype $x$ produces the Polygenetic Score (PGS) as output,

$$\text{PGS} = x \cdot \beta\,. \tag{2}$$

The LASSO objective function (1) favors sparsity, as controlled by $\lambda$, making it suitable for genetic prediction [82]. A well-chosen $\lambda$ allows the predictor to focus on the comparably few relevant SNPs among the $\sim$ 50k used as input as selected by GWAS p-value.

For this work, LASSO was implemented using a standard package of Python3 [83]: Scikit-Learn [84]. Additionally, we used a custom implementation of LASSO, developed in Julia, as a cross check. Further details about this custom implementation can be found in [11,42,43]. All code was run on the Michigan State University High Performance Computing Cluster (HPCC). With parallelization, predictors can be trained on $\sim 300k - 500k$ samples in $\sim 12 - 36$ hours. Genotype input, $X$, was prepared using PLINK[85,86] and read with PySNPTools [87].

Within a population we can classify how much predicted variance there is as

$$\begin{aligned} \text{var}\left(x \cdot \beta\right) &= \sum_i \left( \text{var}\left(x_i \beta_i\right) + 2 \sum_{j<i} \text{cov}\left(x_i \beta_i, x_j \beta_j\right) \right) \\ &= \sum_i \left( 2\beta_i^2 (1 - f_i) f_i + 2 \sum_{j<i} \text{cov}\left(x_i \beta_i, x_j \beta_j\right) \right)\,. \end{aligned} \tag{3}$$

The variance accounted for *from single SNPs alone* can be described as

$$\sum_i 2\beta_i^2 (1 - f_i) f_i\,. \tag{4}$$

This is equivalent to the *total* variance of the predictor when correlations among the SNPs are small. Sparse prediction methods generally suppress activating SNPs which are highly correlated, reducing the impact of the neglected covariance. Further details can be found in [42].

#### S2 Phenotype Descriptions

The research underlying this paper was conducted over the course of two data updates to a UKB application. The first dataset was acquired from UKB in April 2019 and was used for all the PGS research. The second, mostly overlapping, dataset was acquired in November 2020 and was used for the disease prediction research. The phenotype availability differed for these datasets why the sample sizes are not the same for the PGS-related results as for the (risk score | biomarkers) results.

##### S2.1 Phenotype distribution by sex for European ancestry

**Figure S1** and **Figure S2** show phenotype distributions for both females and males of European ancestry with the number of samples being used for the PGS training shown in the upper corner of each plot.

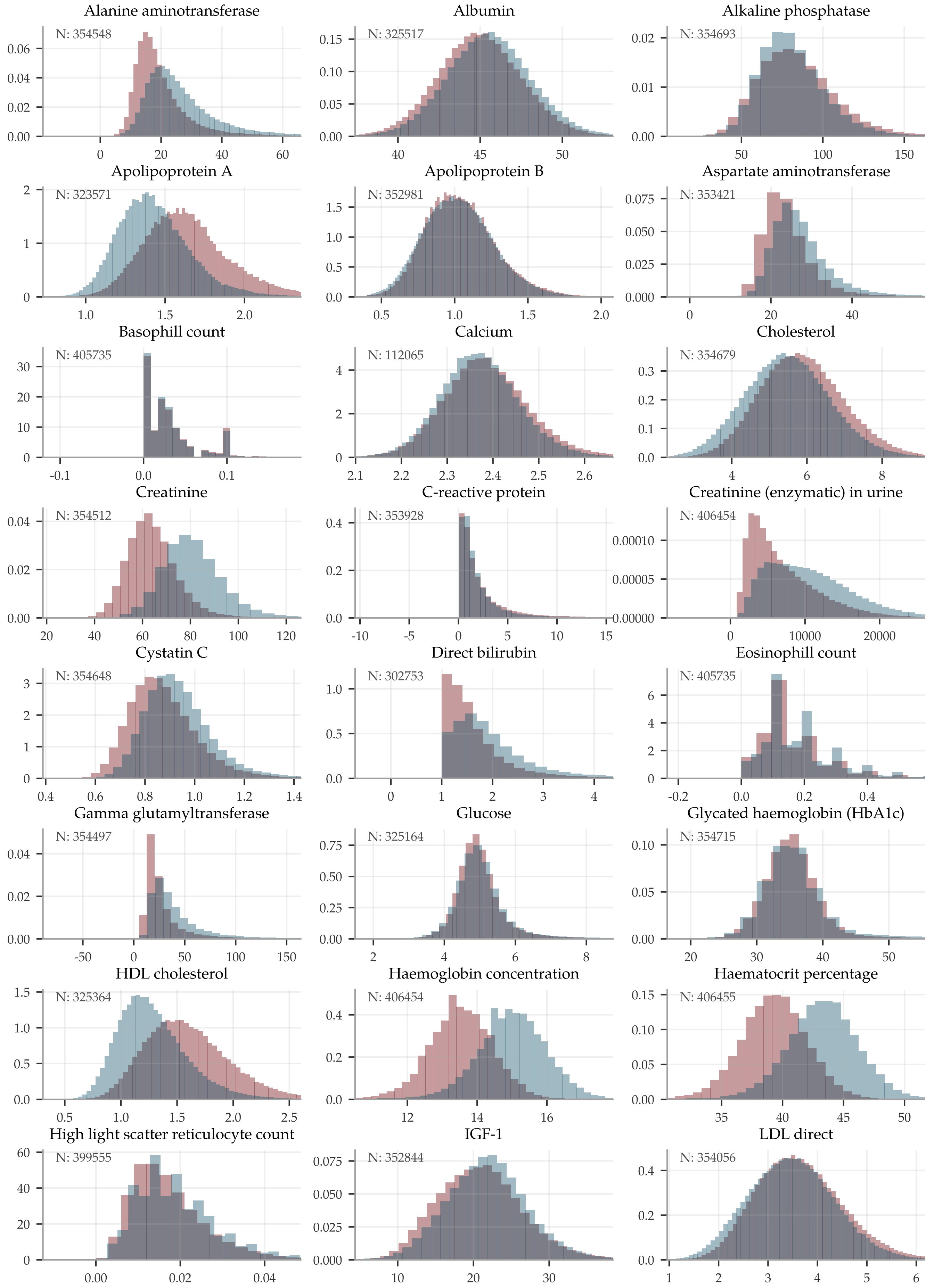


**Figure S1: Distribution plots (1 of 2) for European ■ men and ■ women of the raw biomarker measurements.** The x-axis are cropping some of the longer tails.

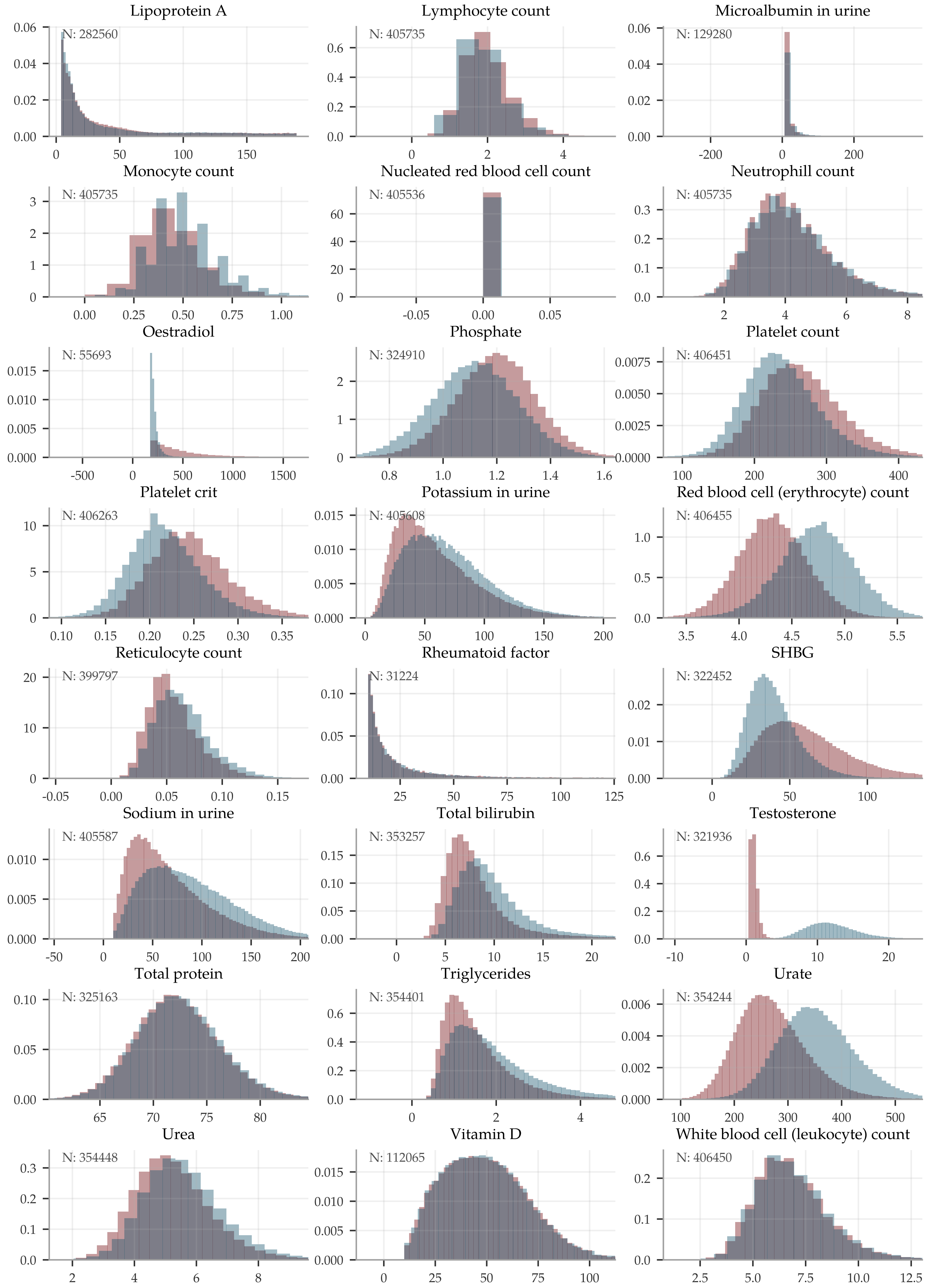


**Figure S2: Distribution plots (2 of 2) for European ■ men and ■ women of the raw biomarker measurements.** The x-axis are cropping some of the longer tails.

#### S2.2 Phenotype dependence on age

All individuals of European ancestry where binned by age at biomarker measurement and the normalized means in each bin are showed in **Figure S3**.

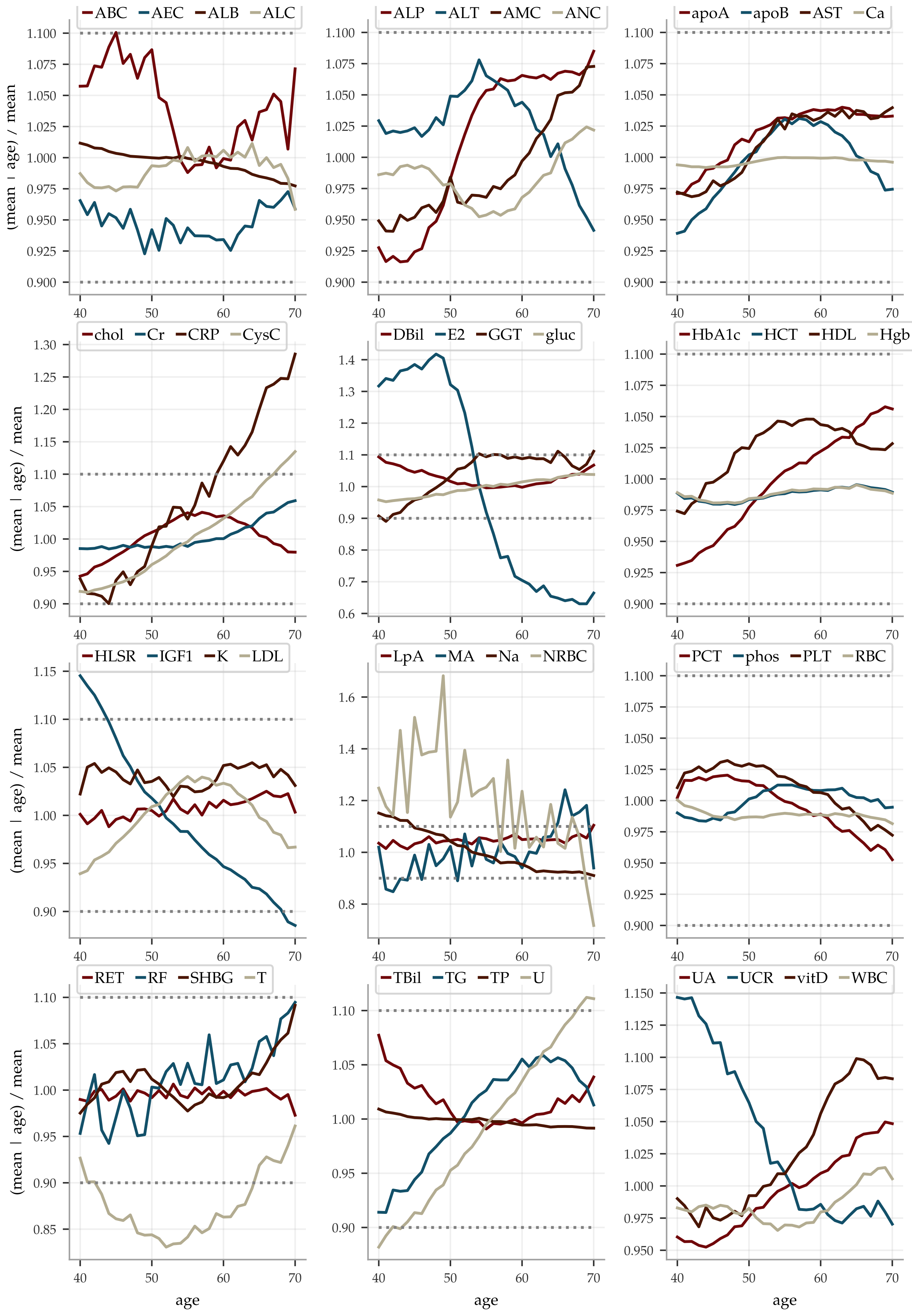


**Figure S3: Age dependence of each of the biomarkers.** The binned means of all Europeans of the same age are shown, normalized with the overall mean.

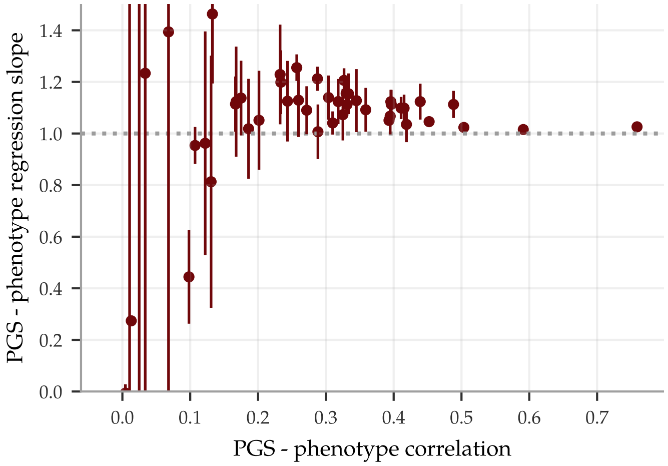


**Figure S4: Calibration of PGS predictors is poor for predictors with low correlation and stabilizes somewhat for those predictors with correlations over 0.3.** The error bars indicate the standard deviation for the five predictor for each trait. The numbers are shown in **Table S1**.

| Abbr. | Slope ± std | Abbr. | Slope ± std | Abbr. | Slope ± std |
|---|---|---|---|---|---|
| ABC | 1.46 ± 0.27 | DBil | 1.02 ± 0.00 | PCT | 1.12 ± 0.07 |
| AEC | 1.14 ± 0.09 | E2 | -0.05 ± 0.03 | phos | 1.13 ± 0.14 |
| ALB | 1.21 ± 0.05 | GGT | 1.20 ± 0.12 | PLT | 1.11 ± 0.05 |
| ALC | 0.95 ± 0.07 | gluc | 0.96 ± 0.43 | RBC | 1.04 ± 0.07 |
| ALP | 1.10 ± 0.04 | HbA1c | 1.01 ± 0.11 | RET | 1.12 ± 0.21 |
| ALT | 1.05 ± 0.19 | HCT | 1.15 ± 0.08 | RF | 1.23 ± 2.30 |
| AMC | 1.09 ± 0.09 | HDL | 1.05 ± 0.02 | SHBG | 1.07 ± 0.07 |
| ANC | 1.12 ± 0.09 | Hgb | 1.13 ± 0.12 | T | 1.11 ± 0.11 |
| apoA | 1.10 ± 0.05 | HLSR | 1.16 ± 0.05 | TBil | 1.02 ± 0.01 |
| apoB | 1.05 ± 0.02 | IGF1 | 1.11 ± 0.06 | TG | 1.09 ± 0.08 |
| AST | 1.02 ± 0.19 | K | 1.39 ± 1.56 | TP | 1.21 ± 0.05 |
| Ca | 5.68 ± 10.35 | LDL | 1.11 ± 0.03 | U | 1.23 ± 0.19 |
| chol | 1.07 ± 0.10 | LpA | 1.03 ± 0.00 | UA | 1.12 ± 0.05 |
| Cr | 1.13 ± 0.16 | MA | NAN | UCR | 0.44 ± 0.18 |
| CRP | 1.14 ± 0.14 | Na | 0.81 ± 0.49 | vitD | 0.27 ± 0.01 |
| CysC | 1.04 ± 0.04 | NRBC | -0.01 ± 0.03 | WBC | 1.26 ± 0.05 |

**Table S1: Calibration of the PGS predictors.**

### S3 Phenotype Prediction

#### S3.1 Calibration

The calibration, i.e. the slope of a linear regression of the PGS predictors, varied: the poorly performing predictors clearly failed the calibration tests while all PGS with a correlation above 0.3 fared better. The calibration for each biomarker is plotted in **Figure S4** against the PGS-phenotype correlation and the numerical values are listed in **Table S1**.

#### S3.2 PGS correlations

The numerical values for the PGS-phenotype correlations are listed in **Table S2** as means ± the standard deviations for 5 predictors trained on each biomarker.

| | European | | South Asian | | East Asian | | African | |
|---|---|---|---|---|---|---|---|---|
| Abbr. | N | Corr | N | Corr | N | Corr | N | Corr |
| ABC | 405735/38885 | 0.127 ± 0.011 | 9070 | 0.103 ± 0.003 | 1453 | 0.009 ± 0.046 | 7221 | -0.006 ± 0.000 |
| AEC | 405735/38885 | 0.302 ± 0.002 | 9070 | 0.250 ± 0.002 | 1453 | 0.172 ± 0.010 | 7221 | 0.089 ± 0.011 |
| ALB | 325517/35240 | 0.287 ± 0.001 | 8217 | 0.244 ± 0.001 | 1296 | 0.217 ± 0.004 | 6632 | 0.123 ± 0.004 |
| ALC | 405735/38885 | 0.107 ± 0.001 | 9070 | 0.121 ± 0.000 | 1453 | 0.136 ± 0.002 | 7221 | 0.012 ± 0.002 |
| ALP | 354693/38326 | 0.409 ± 0.001 | 8989 | 0.390 ± 0.005 | 1422 | 0.277 ± 0.013 | 7170 | 0.115 ± 0.010 |
| ALT | 354548/38320 | 0.196 ± 0.003 | 8973 | 0.159 ± 0.010 | 1421 | 0.135 ± 0.019 | 7168 | 0.027 ± 0.006 |
| AMC | 405735/38885 | 0.269 ± 0.002 | 9070 | 0.248 ± 0.005 | 1453 | 0.239 ± 0.016 | 7221 | 0.018 ± 0.004 |
| ANC | 405735/38885 | 0.316 ± 0.001 | 9070 | 0.266 ± 0.004 | 1453 | 0.182 ± 0.007 | 7221 | 0.206 ± 0.023 |
| apoA | 323571/35052 | 0.413 ± 0.003 | 8201 | 0.364 ± 0.006 | 1284 | 0.328 ± 0.012 | 6603 | 0.218 ± 0.016 |
| apoB | 352981/38157 | 0.393 ± 0.001 | 8906 | 0.254 ± 0.003 | 1413 | 0.282 ± 0.005 | 7107 | 0.243 ± 0.006 |
| AST | 353421/38193 | 0.181 ± 0.003 | 8956 | 0.127 ± 0.009 | 1410 | 0.055 ± 0.001 | 7131 | 0.014 ± 0.001 |
| Ca | 112065/11643 | 0.015 ± 0.006 | 2188 | 0.001 ± 0.012 | 413 | 0.042 ± 0.041 | 1892 | 0.013 ± 0.010 |
| chol | 354679/38321 | 0.321 ± 0.006 | 8988 | 0.202 ± 0.013 | 1422 | 0.245 ± 0.021 | 7166 | 0.181 ± 0.030 |
| Cr | 354512/38298 | 0.240 ± 0.001 | 8987 | 0.074 ± 0.004 | 1421 | 0.011 ± 0.001 | 7164 | 0.014 ± 0.004 |
| CRP | 353928/38258 | 0.170 ± 0.003 | 8957 | 0.189 ± 0.005 | 1418 | 0.139 ± 0.011 | 7156 | 0.053 ± 0.011 |
| CysC | 354648/38326 | 0.309 ± 0.001 | 8989 | 0.146 ± 0.002 | 1423 | 0.051 ± 0.001 | 7166 | 0.046 ± 0.003 |
| DBil | 302753/32472 | 0.503 ± 0.000 | 7464 | 0.428 ± 0.000 | 1213 | 0.303 ± 0.003 | 5937 | 0.284 ± 0.008 |
| E2 | 55693/5897 | -0.015 ± 0.009 | 1629 | 0.013 ± 0.008 | 317 | 0.014 ± 0.027 | 1880 | 0.025 ± 0.018 |
| GGT | 354497/38309 | 0.229 ± 0.004 | 8987 | 0.198 ± 0.010 | 1421 | 0.126 ± 0.024 | 7164 | 0.040 ± 0.012 |
| gluc | 325164/35200 | 0.112 ± 0.008 | 8216 | 0.060 ± 0.020 | 1298 | 0.090 ± 0.016 | 6623 | 0.025 ± 0.026 |
| HbA1c | 354715/38306 | 0.286 ± 0.004 | 8885 | 0.169 ± 0.013 | 1428 | 0.160 ± 0.009 | 6219 | 0.024 ± 0.005 |
| HCT | 406455/38943 | 0.332 ± 0.001 | 9102 | 0.204 ± 0.003 | 1453 | 0.196 ± 0.009 | 7243 | 0.115 ± 0.005 |
| HDL | 325364/35234 | 0.451 ± 0.001 | 8212 | 0.372 ± 0.002 | 1298 | 0.305 ± 0.004 | 6630 | 0.240 ± 0.005 |
| Hgb | 406454/38943 | 0.341 ± 0.003 | 9102 | 0.188 ± 0.003 | 1453 | 0.187 ± 0.013 | 7243 | 0.117 ± 0.006 |
| HLSR | 399555/38356 | 0.328 ± 0.003 | 8867 | 0.281 ± 0.002 | 1422 | 0.271 ± 0.005 | 7051 | 0.125 ± 0.006 |
| IGF1 | 352844/38105 | 0.395 ± 0.001 | 8931 | 0.325 ± 0.005 | 1415 | 0.255 ± 0.004 | 7118 | 0.196 ± 0.009 |
| K | 405608/38906 | 0.054 ± 0.016 | 9036 | 0.029 ± 0.015 | 1443 | 0.032 ± 0.014 | 7332 | 0.007 ± 0.012 |
| LDL | 354056/38261 | 0.330 ± 0.002 | 8969 | 0.203 ± 0.002 | 1420 | 0.249 ± 0.006 | 7152 | 0.225 ± 0.009 |
| LpA | 282560/30694 | 0.759 ± 0.000 | 7889 | 0.351 ± 0.001 | 1282 | 0.110 ± 0.001 | 5995 | 0.088 ± 0.000 |
| MA | 129280/11918 | 0.002 ± 0.008 | 3284 | -0.003 ± 0.003 | 427 | 0.011 ± 0.034 | 3372 | 0.015 ± 0.002 |
| Na | 405587/38912 | 0.122 ± 0.006 | 9035 | 0.084 ± 0.004 | 1444 | -0.003 ± 0.018 | 7328 | 0.035 ± 0.004 |
| NRBC | 405536/38864 | 0.001 ± 0.003 | 9070 | -0.002 ± 0.004 | 1453 | 0.022 ± 0.025 | 7221 | 0.025 ± 0.003 |
| PCT | 406263/38923 | 0.437 ± 0.003 | 9101 | 0.365 ± 0.004 | 1453 | 0.292 ± 0.011 | 7243 | 0.152 ± 0.009 |
| phos | 324910/35172 | 0.255 ± 0.002 | 8207 | 0.226 ± 0.009 | 1296 | 0.199 ± 0.020 | 6621 | 0.112 ± 0.013 |
| PLT | 406451/38943 | 0.486 ± 0.001 | 9102 | 0.416 ± 0.002 | 1453 | 0.343 ± 0.000 | 7243 | 0.219 ± 0.007 |
| RBC | 406455/38943 | 0.417 ± 0.002 | 9102 | 0.277 ± 0.004 | 1453 | 0.227 ± 0.010 | 7243 | 0.161 ± 0.006 |
| RET | 399797/38388 | 0.162 ± 0.002 | 8869 | 0.167 ± 0.018 | 1423 | 0.199 ± 0.028 | 7051 | 0.050 ± 0.001 |
| RF | 31224/3336 | 0.033 ± 0.013 | 737 | 0.046 ± 0.018 | 112 | 0.038 ± 0.038 | 534 | -0.012 ± 0.022 |
| SHBG | 322452/34921 | 0.392 ± 0.002 | 8162 | 0.289 ± 0.005 | 1288 | 0.255 ± 0.001 | 6567 | 0.173 ± 0.019 |
| T | 321936/34635 | 0.164 ± 0.003 | 8217 | 0.140 ± 0.002 | 1329 | 0.069 ± 0.005 | 6495 | 0.017 ± 0.009 |
| TBil | 353257/38169 | 0.591 ± 0.000 | 8951 | 0.554 ± 0.001 | 1409 | 0.473 ± 0.003 | 7126 | 0.431 ± 0.018 |
| TG | 354401/38292 | 0.356 ± 0.004 | 8977 | 0.310 ± 0.013 | 1421 | 0.242 ± 0.013 | 7164 | 0.127 ± 0.019 |
| TP | 325163/35189 | 0.326 ± 0.001 | 8218 | 0.278 ± 0.001 | 1297 | 0.244 ± 0.006 | 6631 | 0.118 ± 0.007 |
| U | 354448/38292 | 0.228 ± 0.004 | 8987 | 0.165 ± 0.003 | 1420 | 0.101 ± 0.006 | 7164 | 0.059 ± 0.012 |
| UA | 354244/38288 | 0.393 ± 0.003 | 8977 | 0.328 ± 0.001 | 1420 | 0.287 ± 0.005 | 7160 | 0.204 ± 0.017 |
| UCR | 406454/38985 | 0.097 ± 0.005 | 9051 | 0.049 ± 0.002 | 1445 | 0.031 ± 0.005 | 7335 | 0.027 ± 0.005 |
| vitD | 112065/11643 | 0.013 ± 0.000 | 2188 | 0.016 ± 0.004 | 413 | 0.046 ± 0.012 | 1892 | -0.024 ± 0.003 |
| WBC | 406450/38943 | 0.256 ± 0.001 | 9102 | 0.256 ± 0.001 | 1453 | 0.201 ± 0.002 | 7243 | 0.077 ± 0.002 |

**Table S2: PGS correlations for both sexes broken down according to ancestry.** N denotes evaluation set sizes and training/evaluation set sizes in the case of European ancestry.

##### S3.3 Sibling evaluation

The main document presented the sibling evaluation only for the PGS predictors with the highest correlations. The full figure with all biomarkers is shown in **Figure S5**, while the sample sizes used are listed in **Table S3** (vitamin D was left out of this analysis due to too few samples). Again, sib 0.5, sib 1.0 and sib 1.5, are the results when restricting to siblings with phenotype differences larger than 0.5, 1 and 1.5 standard deviations, respectively.

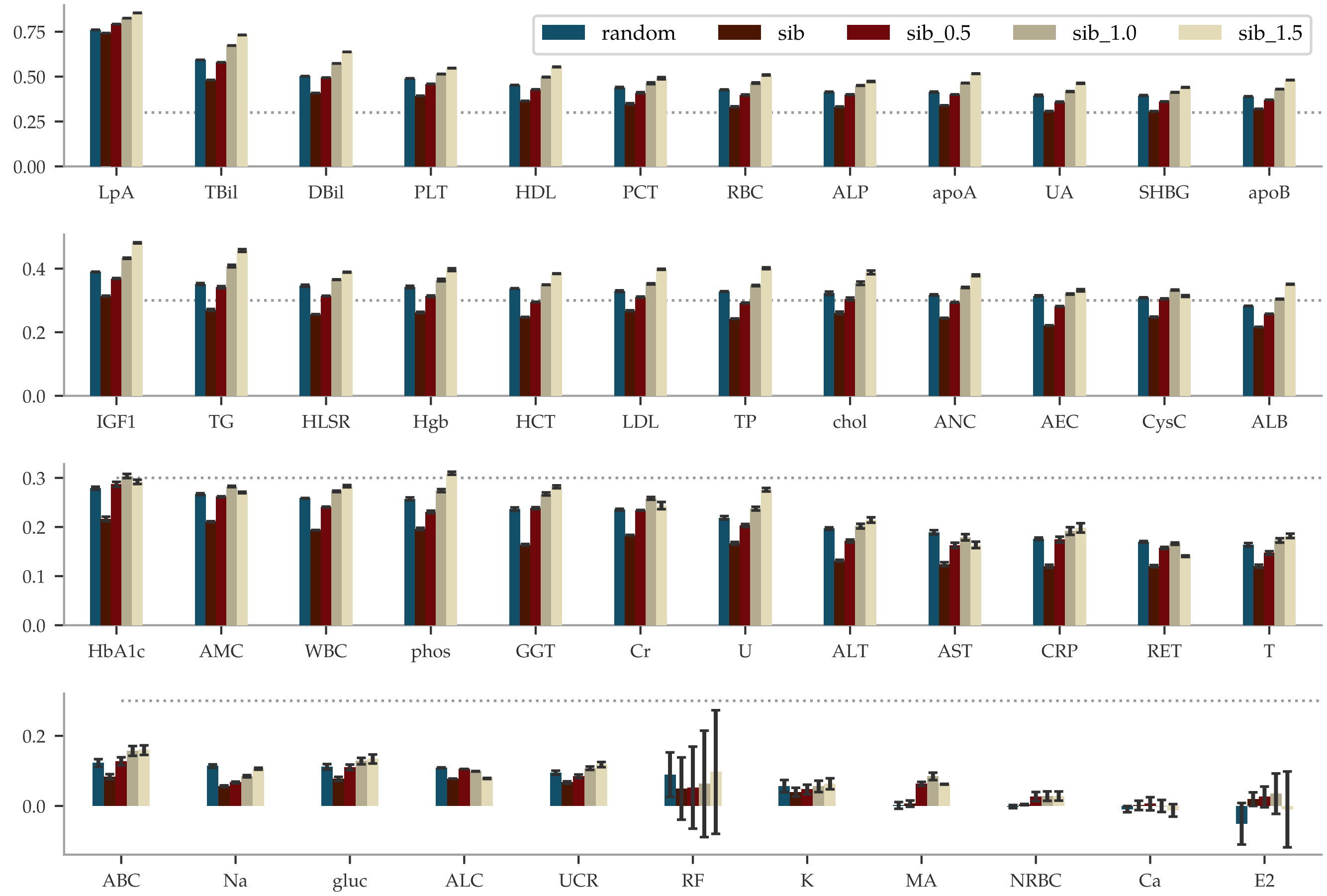


**Figure S5: Sibling comparisons of correlation between difference in phenotype and difference in PGS, i.e.,** $\mathrm{corr}(\Delta_{\mathrm{phen}}, \Delta_{\mathrm{PGS}})$**.** UKBs $\tilde{4}0$k European siblings were paired either randomly or as genetic siblings. The error bars indicate ± the standard deviations for 5 predictors trained on slightly different training sets. The additional three bars labeled sib 0.5, sib 1.0 and sib 1.5, are the results when restricting to siblings with phenotype differences larger than 0.5, 1 and 1.5 standard deviations, respectively. Vitamin D was left out because of too few samples.

| Abbr. | random | sib | sib_0.5 | sib_1.0 | sib_1.5 | Abbr. | random | sib | sib_0.5 | sib_1.0 | sib_1.5 |
|---|---|---|---|---|---|---|---|---|---|---|---|
| ABC | 19638 | 20469 | 8442 | 4631 | 2992 | HLSR | 19109 | 19947 | 11704 | 6032 | 2954 |
| AEC | 19638 | 20469 | 10512 | 4714 | 2189 | IGF1 | 18873 | 19672 | 12913 | 7297 | 3673 |
| ALB | 16159 | 16861 | 11401 | 6889 | 3668 | K | 19626 | 20465 | 14023 | 8984 | 5416 |
| ALC | 19638 | 20469 | 6964 | 1623 | 403 | LDL | 19014 | 19829 | 13478 | 8319 | 4589 |
| ALP | 19086 | 19897 | 11675 | 5830 | 2737 | LpA | 12203 | 13386 | 4967 | 2921 | 2116 |
| ALT | 19078 | 19890 | 10125 | 4904 | 2617 | MA | 1829 | 2173 | 187 | 99 | 60 |
| AMC | 19638 | 20469 | 10748 | 4660 | 1843 | Na | 19636 | 20474 | 13260 | 7799 | 4321 |
| ANC | 19638 | 20469 | 13186 | 7498 | 3938 | NRBC | 19613 | 20445 | 427 | 416 | 383 |
| apoA | 15980 | 16679 | 11047 | 6502 | 3440 | PCT | 19672 | 20504 | 12949 | 7130 | 3518 |
| apoB | 18918 | 19723 | 13301 | 8078 | 4421 | phos | 16093 | 16792 | 11540 | 7164 | 3984 |
| AST | 18952 | 19767 | 9259 | 3804 | 1715 | PLT | 19696 | 20527 | 12907 | 7035 | 3426 |
| Ca | 1756 | 2473 | 1493 | 763 | 396 | RBC | 19696 | 20527 | 12783 | 6899 | 3174 |
| chol | 19082 | 19893 | 13501 | 8184 | 4490 | RET | 19144 | 19983 | 8492 | 2666 | 715 |
| Cr | 19054 | 19865 | 9613 | 3531 | 1266 | RF | 155 | 223 | 120 | 73 | 48 |
| CRP | 19016 | 19825 | 6367 | 3174 | 1964 | SHBG | 15874 | 16552 | 10487 | 5881 | 3155 |
| CysC | 19087 | 19897 | 10476 | 4506 | 1852 | T | 15598 | 16326 | 10503 | 6015 | 3180 |
| DBil | 13657 | 14459 | 7912 | 4107 | 2208 | TBil | 18921 | 19738 | 10334 | 4891 | 2550 |
| E2 | 446 | 774 | 383 | 205 | 122 | TG | 19045 | 19860 | 11203 | 6170 | 3431 |
| GGT | 19065 | 19877 | 6538 | 3148 | 1832 | TP | 16116 | 16810 | 10919 | 6198 | 3203 |
| gluc | 16117 | 16819 | 6466 | 2435 | 1256 | U | 19046 | 19853 | 12931 | 7440 | 3797 |
| HbA1c | 19060 | 19886 | 8339 | 2901 | 1334 | UA | 19048 | 19858 | 13164 | 7703 | 4034 |
| HCT | 19696 | 20527 | 13477 | 7831 | 3944 | UCR | 19714 | 20555 | 12872 | 7594 | 4194 |
| HDL | 16151 | 16853 | 11004 | 6384 | 3318 | WBC | 19696 | 20527 | 11937 | 5693 | 2424 |
| Hgb | 19696 | 20527 | 13103 | 7259 | 3609 | | | | | | |

**Table S3: Number of pairs in each evaluation set used for Figure S5.**

#### S3.4 Genetic Architectures

The $\beta$ coefficient sizes of one predictor for every biomarker are plotted in **Figure S6** to **Figure S11**. The same figures also feature the cumulative variance accounted for from single SNPs, as described in eq. (4). Number of non-zero coefficients ($\pm$ standard deviation over 5 predictors) and PGS-phenotype correlations are listed within each plot title.

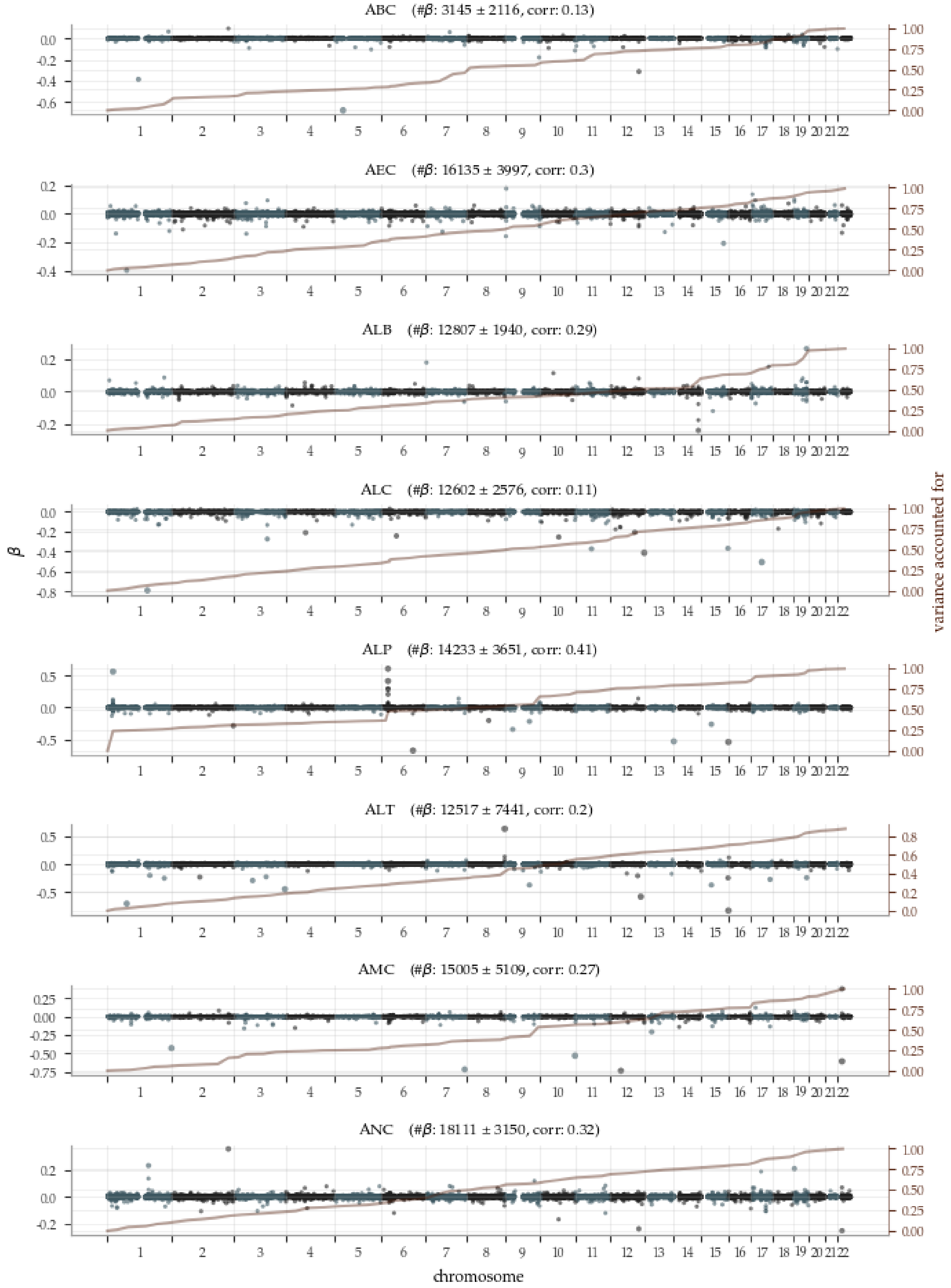


**Figure S6:** Manhattan plots (1 of 6) of PGS predictor $\beta$ with superimposed variance accounted for.

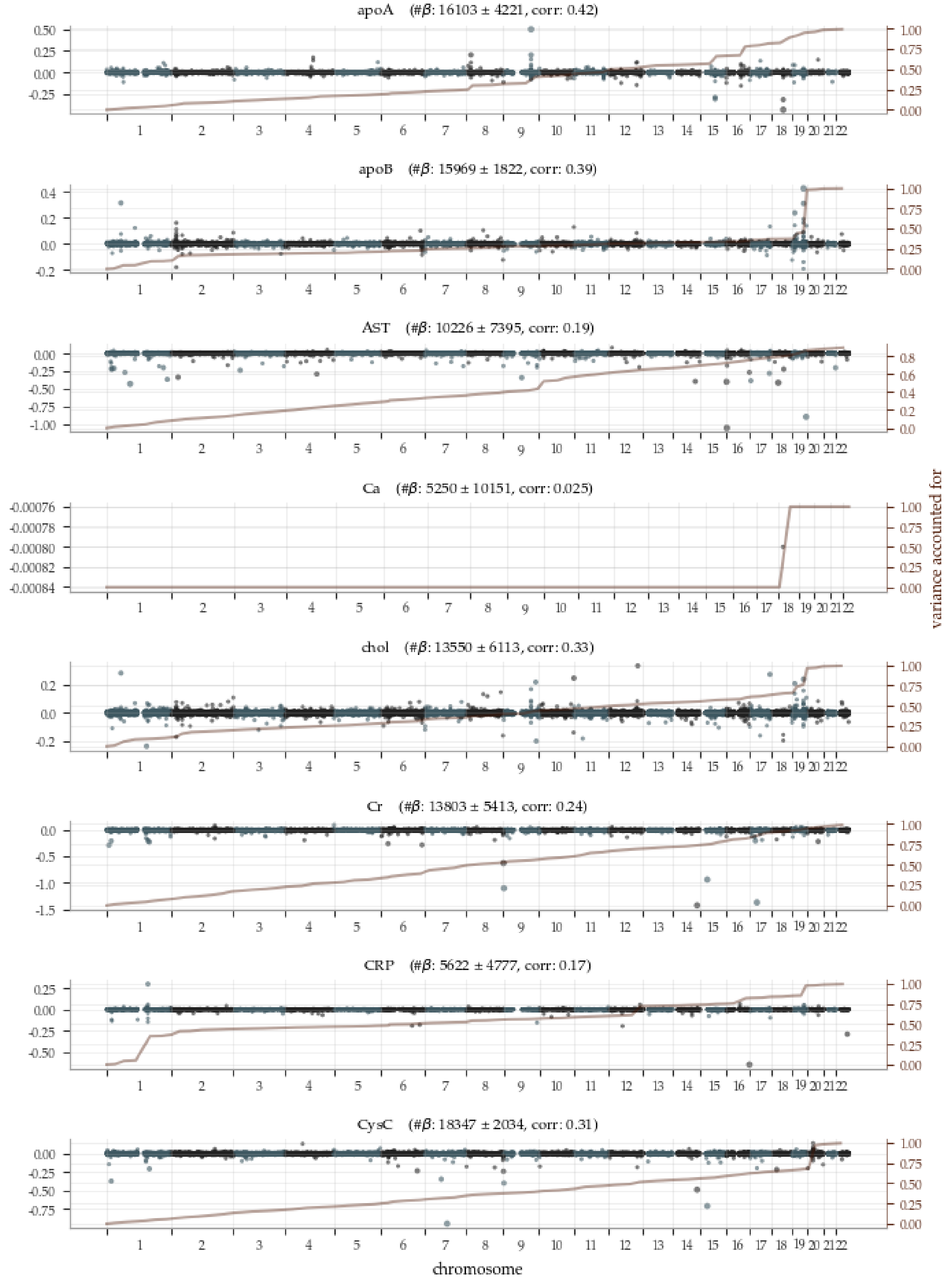


**Figure S7:** Manhattan plots (2 of 6) of PGS predictor $\beta$ with superimposed variance accounted for.

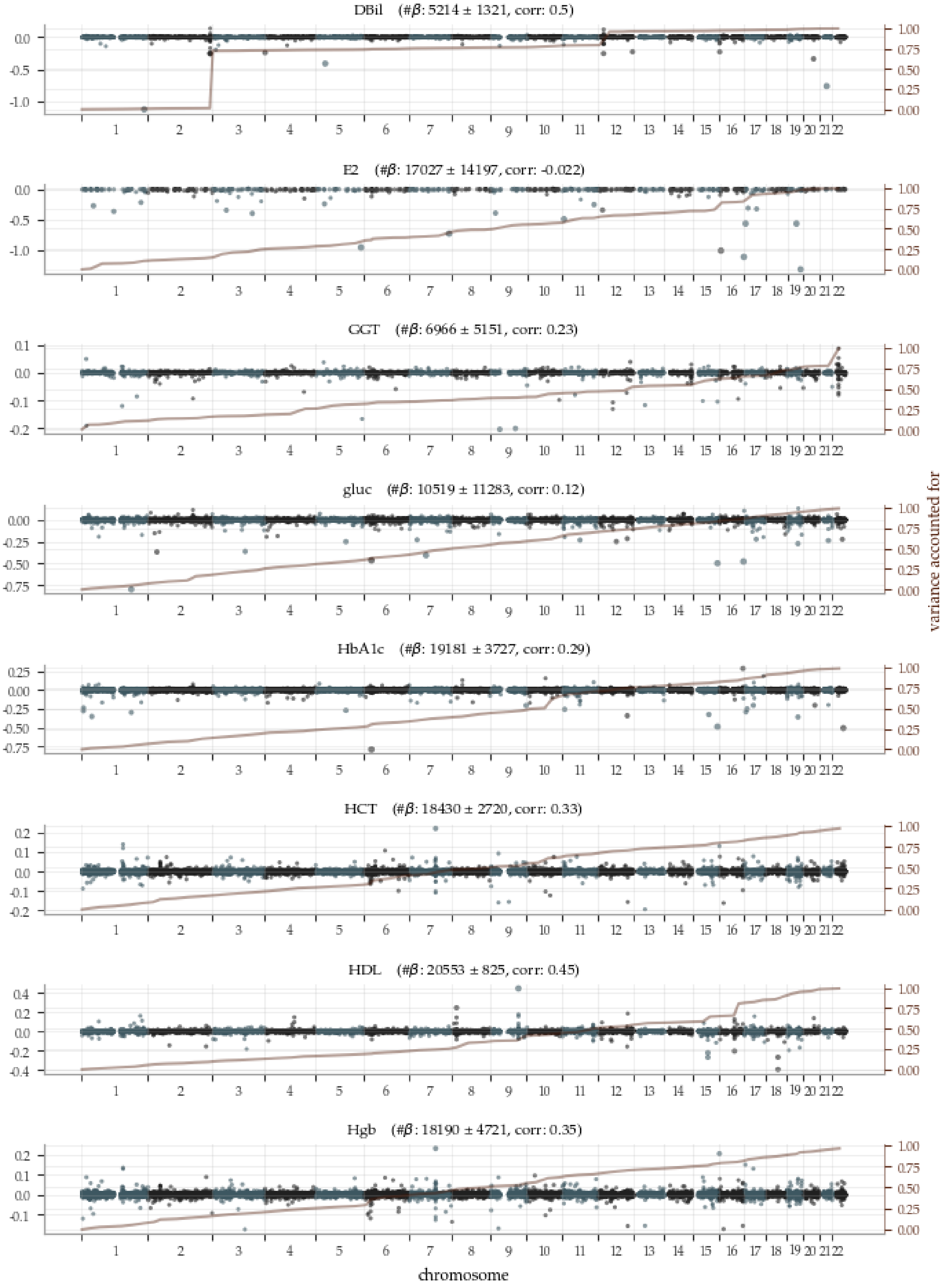


**Figure S8:** Manhattan plots (3 of 6) of PGS predictor $\beta$ with superimposed variance accounted for.

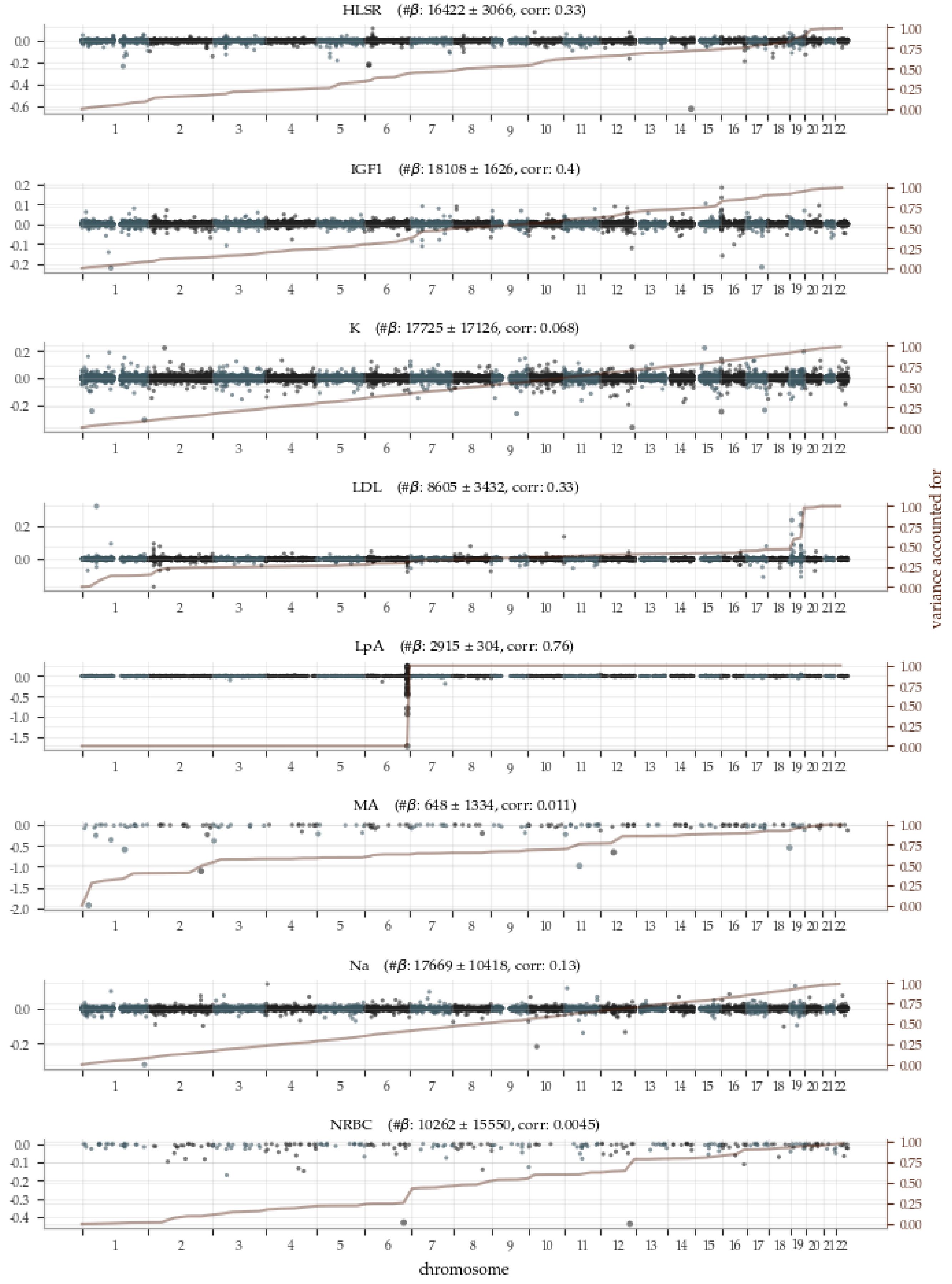


**Figure S9:** Manhattan plots (4 of 6) of PGS predictor $\beta$ with superimposed variance accounted for.

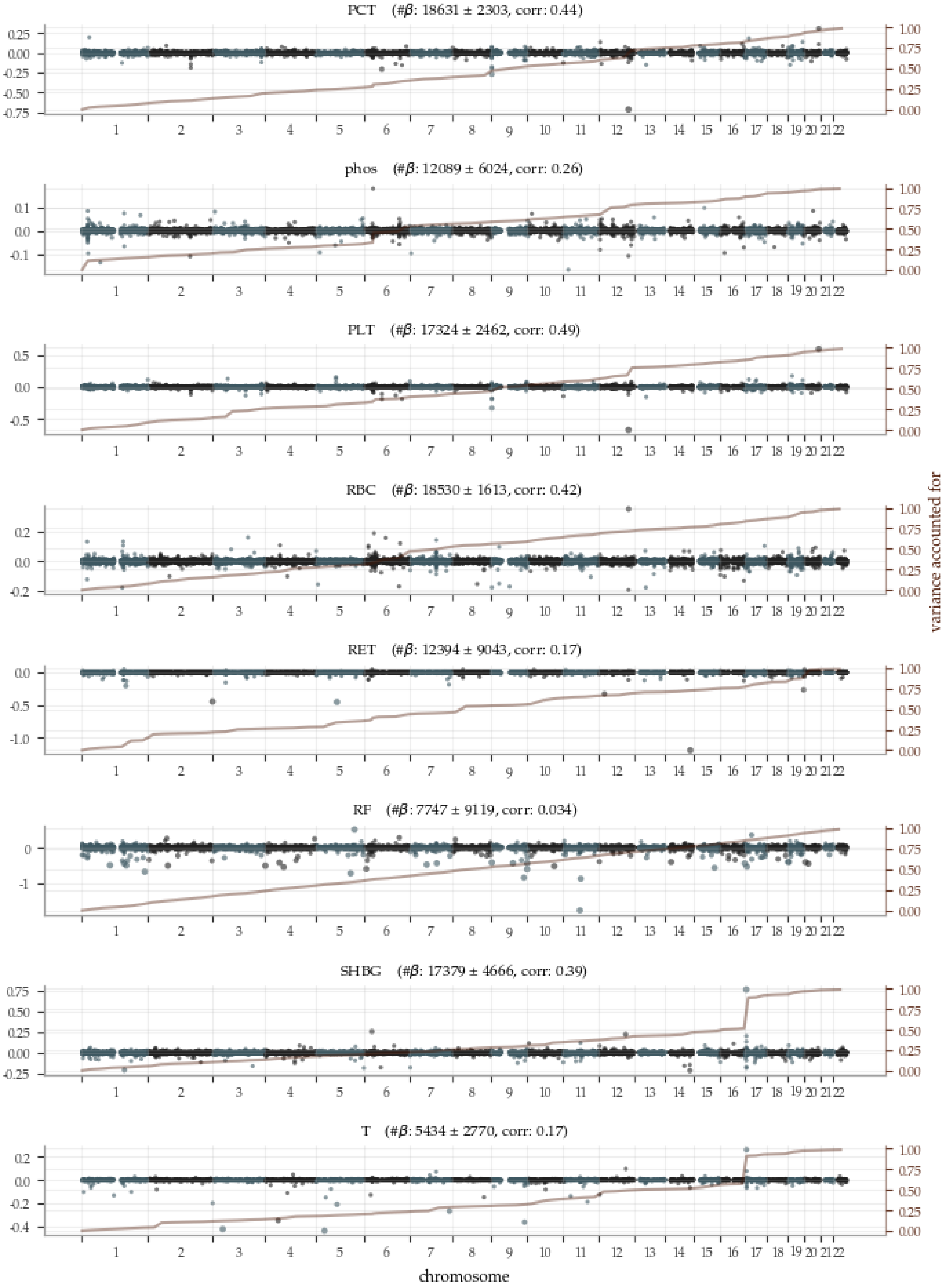


**Figure S10:** Manhattan plots (5 of 6) of PGS predictor $\beta$ with superimposed variance accounted for.

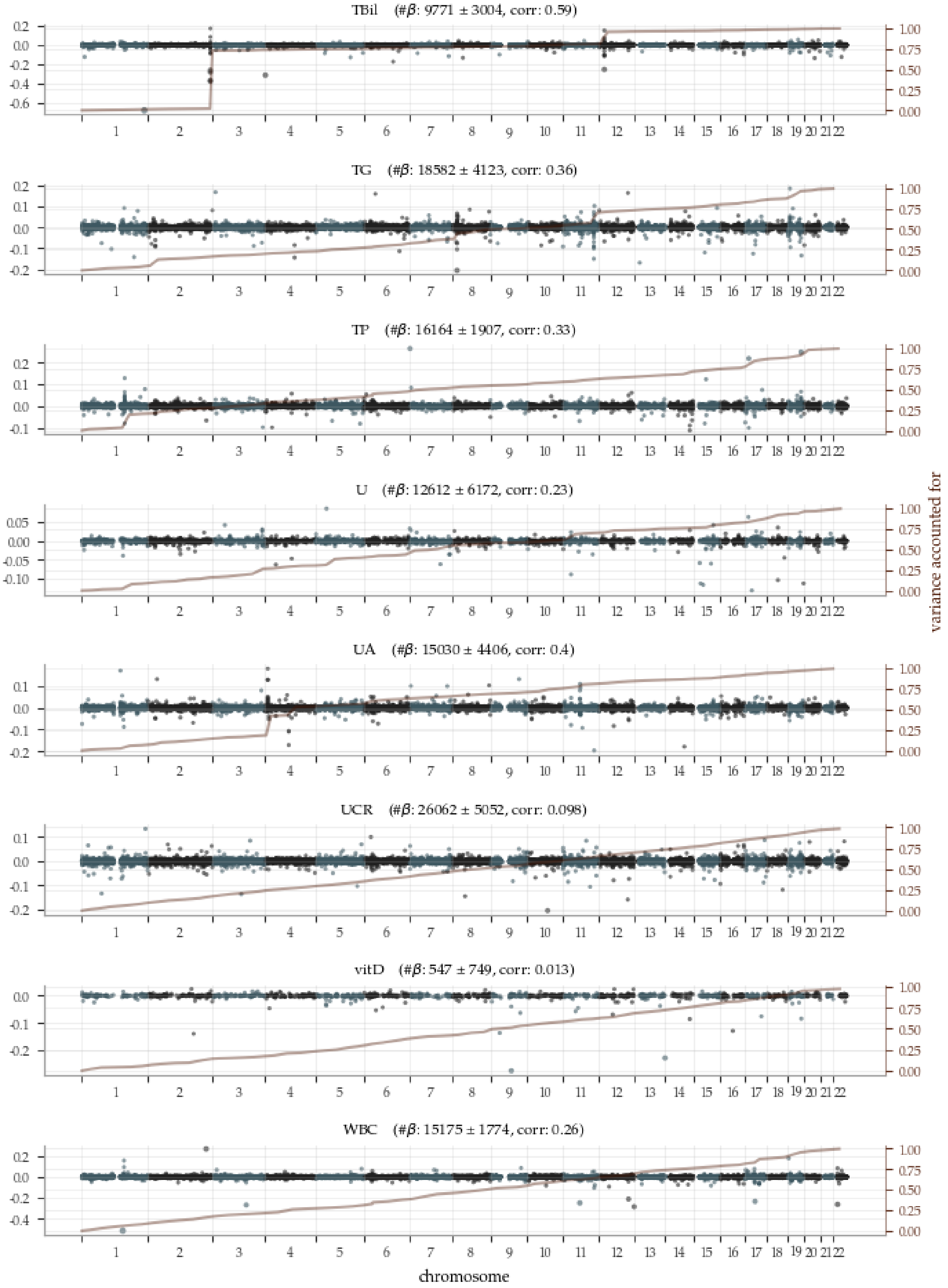


**Figure S11:** Manhattan plots (6 of 6) of PGS predictor $\beta$ with superimposed variance accounted for.

### S4 Disease Prediction

#### S4.1 Disease condition definitions

The disease conditions were defined in terms of matching codes in a number of fields in the UKB. Each condition is listed below with all its codes for all UKB fields involved. An individual was classified as a case if any of the disease codes was present.

##### CAD

**Non-cancer illness code, self-reported, UKB field 20002** 1075

**Diagnoses - ICD9, UKB field 41271** 410, 4109, 412, 4129

**Diagnoses - ICD10, UKB field 41270** I210, I211, I212, I213, I214, I219, I21X, I22, I220, I221, I228, I229, I23, I230, I231, I232, I233, I234, I235, I236, I238, I241, I252

**Operative procedures - OPCS4, UKB field 41272** K401, K402, K403, K404, K411, K412, K413, K414, K451, K452, K453, K454, K455, K491, K492, K498, K499, K502, K751, K752, K753, K754, K758, K759

##### Cancer

**Cancer code, self-reported, UKB field 20001** -1, 1001, 1002, 1003, 1004, 1005, 1006, 1007, 1008, 1009, 1010, 1011, 1012, 1015, 1016, 1017, 1018, 1019, 1020, 1021, 1022, 1023, 1024, 1025, 1026, 1027, 1028, 1029, 1030, 1031, 1032, 1033, 1034, 1035, 1036, 1037, 1038, 1039, 1040, 1041, 1042, 1043, 1044, 1045, 1046, 1047, 1048, 1050, 1051, 1052, 1053, 1055, 1056, 1058, 1059, 1060, 1061, 1062, 1063, 1064, 1065, 1066, 1067, 1068, 1070, 1071, 1072, 1073, 1074, 1075, 1076, 1077, 1078, 1079, 1080, 1081, 1082, 1084, 1085, 1086, 1087, 1088, 99999

##### Diabetes (irrespective of type, used in ASCVD Risk Estimator input)

**Non-cancer illness code, self-reported, UKB field 20002** 1220, 1222, 1223

**Diagnoses - ICD9, UKB field 41271** 250, 2500, 25000, 25001, 25009, 2501, 25010, 25011, 25019, 2502, 25020, 25021, 25029, 2503, 2504, 2505, 2506, 2507, 2509, 25090, 25091, 25099, 3572, 3620, 7751

**Diagnoses - ICD10, UKB field 41270** E10, E100, E101, E102, E103, E104, E105, E106, E107, E108, E109, E11, E110, E111, E112, E113, E114, E115, E116, E117, E118, E119, E12, E120, E121, E122, E123, E124, E125, E126, E127, E128, E129, E13, E130, E131, E132, E133, E134, E135, E136, E137, E138, E139, E14, E140, E141, E142, E143, E144, E145, E146, E147, E148, E149, G590, G632, H280, H360, M142, N083, O240, O241, O242, O243, P702

##### Diabetes type 1

**Non-cancer illness code, self-reported, UKB field 20002** 1220

**Diagnoses - ICD9, UKB field 41271** 25001, 25003, 25011, 25013, 25021, 25023, 25031, 25033, 25041, 25043, 25051, 25053, 25061, 25063, 25071, 25073, 25081, 25083, 25091, 25093

**Diagnoses - ICD10, UKB field 41270** E10, E100, E101, E102, E103, E104, E105, E106, E107, E108, E109

##### Diabetes type 2

**Non-cancer illness code, self-reported, UKB field 20002** 1223

**Diagnoses - ICD9, UKB field 41271** 25000, 25002, 25010, 25012, 25020, 25022, 25030, 25032, 25040, 25042, 25050, 25052, 25060, 25062, 25070, 25072, 25080, 25082, 25090, 25092

**Diagnoses - ICD10, UKB field 41270** E11, E110, E111, E112, E113, E114, E115, E116, E117, E118, E119

##### Hypertension

**Non-cancer illness code, self-reported, UKB field 20002** 1065, 1072

**Diagnoses - ICD9, UKB field 41271** 401, 4010, 4011, 4019, 402, 4020, 4021, 4029, 403, 4030, 4031, 4039, 404, 4040, 4041, 4049, 405, 4050, 4051, 4059, 4160, 4372, 5723

**Diagnoses - ICD10, UKB field 41270** I10, I11, I110, I119, I12, I120, I129, I13, I130, I131, I132, I139, I15, I150, I151, I152, I158, I159, I270, I272, I674, K766

#### Kidney problem

**Cancer code, self-reported, UKB field 20001** 1034

**Non-cancer illness code, self-reported, UKB field 20002** 1192, 1197, 1405, 1427, 1519

**Diagnoses - ICD9, UKB field 41271** 189, 1890, 1898, 1980, 223, 2230, 2238, 2239, 5808, 5818, 5828, 5838, 5839, 5848, 589, 5890, 5891, 5899, 590, 5909, 592, 5920, 593, 5931, 5932, 5938, 59389, 5939, 75300, 75301, 7531, 75310, 75311, 75312, 75313, 75318, 7533, 75330, 75331, 75332, 75333, 75334, 75338, 75390, 7944, 866, 8660, 8661, E8702, E8712, E8722, E8742, E8791, V420, V594

**Diagnoses - ICD10, UKB field 41270** C64, C790, N181, N182, N183, N184, N185, N20, N200, N202, N26, N27, N270, N271, N279, N28, N280, N281, N288, N289, N29, N290, N291, N298, Q60, Q61, Q611, Q612, Q613, Q615, Q618, Q619, Q63, Q630, Q631, Q632, Q633, Q638, Q639, R944, S370, S3700, S3701, T861, Y602, Y612, Y622, Y841, Z524, Z905, Z940

**Operative procedures - OPCS3, UKB field 41273** 5611, 564, 5641, 5642, 5643, 565, 566, 5661, 5662, 567, 5671, 5672, 568, 579, 5792, 5794, 5795

**Operative procedures - OPCS4, UKB field 41272** M01, M011, M012, M013, M014, M015, M018, M019, M02, M024, M026, M027, M028, M029, M03, M031, M032, M038, M039, M04, M041, M042, M043, M048, M049, M05, M054, M055, M058, M059, M06, M061, M062, M068, M069, M07, M071, M072, M078, M079, M08, M081, M082, M083, M084, M088, M089, M09, M091, M092, M093, M094, M098, M099, M10, M101, M103, M104, M105, M108, M109, M11, M111, M112, M113, M118, M119, M13, M131, M132, M133, M134, M135, M137, M138, M139, M14, M141, M148, M149, M15, M158, M159, M16, M161, M162, M168, M169, M17, M171, M172, M173, M174, M175, M178, M179, M196, X451

#### Liver problem

**Non-cancer illness code, self-reported, UKB field 20002** 1136, 1158

**Diagnoses - ICD9, UKB field 41271** 070, 0700, 0701, 0702, 0703, 0704, 0705, 0706, 0709, 1220, 1225, 1228, 1530, 155, 1550, 1551, 1552, 156, 1561, 1568, 1977, 2115, 2308, 2353, 27102, 27103, 27761, 4562, 570, 5709, 571, 5714, 5715, 57150, 57151, 57152, 57158, 57159, 5718, 5719, 572, 5720, 5722, 5728, 573, 5730, 5734, 5738, 5739, 74745, 7516, 75160, 75161, 75162, 75165, 75167, 76782, 7742, 77440, 77441, 77448, 7948, 864, 8640, 8641, V427

**Diagnoses - ICD10, UKB field 41270** A064, B15, B150, B159, B16, B160, B161, B162, B169, B17, B170, B171, B172, B178, B179, B18, B180, B181, B182, B188, B189, B19, B190, B199, B251, B581, B670, B675, B678, B942, C183, C22, C220, C221, C223, C224, C227, C229, C240, C787, D015, D134, D135, D376, P150, P353, Q266, Q44, Q446, Q447, R932, R945, S361, S3610, S3611, T864, Z225, Z526, Z944

**Operative procedures - OPCS3, UKB field 41273** 5005, 502, 5021, 509, 5091, 5092, 5093, 5094, 5097

**Operative procedures - OPCS4, UKB field 41272** J01, J011, J012, J013, J014, J015, J018, J019, J02, J023, J024, J025, J028, J029, J03, J031, J032, J033, J034, J035, J038, J039, J04, J041, J042, J043, J048, J049, J05, J051, J052, J053, J058, J059, J06, J061, J062, J068, J069, J07, J071, J072, J073, J074, J078, J079, J08, J081, J083, J088, J089, J10, J101, J103, J104, J105, J106, J107, J108, J109, J11, J111, J112, J113, J114, J115, J116, J117, J118, J119, J12, J121, J122, J123, J124, J125, J126, J127, J128, J129, J13, J131, J132, J138, J139, J14, J141, J142, J148, J149, J15, J151, J152, J153, J158, J159, J16, J161, J162, J168, J169, J275, J29, J291, J292, J293, J294, J298, J299, J311, J312, J401, J471, J472, J473, J474, J485, J486, J505, J507, J761, J77, J771, J778, J779, O301, O341, O342, O343, O344, O345, T876, X43, X438, X439, X863, X864, Z301, Z376, Z396

#### Obesity

Any individual with BMI, UKB field 21001, larger or equal to 30.

#### ASCVD

**Non-cancer illness code, self-reported, UKB field 20002** 1081, 1074, 1082, 1583

**Diagnoses - ICD9, UKB field 41271** 4139, 4401, 413, 4402, 4140, 4119, 4370, 411, 4148, 4408, 4400, 4149, 4371, 4359, 440, 414, 4409, 435

**Diagnoses - ICD10, UKB field 41270** I7011, I20, I700, I7091, M6228, M6224, M6227, I258, I259, M6222, I7021, P910, I708, P294, I250, I7080, M6226, I256, M6223, G458, I70, I24, I672, I64, I209, I7010, I7001, M6221, I7081, I702, I251, I709, I25, I208, I249, M6220, G459, N280, I7000, I7020, I248, G45, I201, M6229, I255, G463, G464, I200, M6225, I701, I7090, I694, M622

**Operative procedures - OPCS4, UKB field 41272** U543

In addition, ASCVD cases also included any sample that indicated angina, heart attack or stroke in the self-reported UKB field 6150.

##### S4.2 Input data to the ASCVD Risk Estimator

The ASCVD Risk Estimator [24] use 13-14 input data fields. For our evaluation, we used the following UKB data for each of the required inputs. The UKB field IDs are noted within parentheses.

**Age:** Time difference from year(34) and month(52) of birth to date of attending assessment center(21003), rounded to full years.

**Sex:** Self-reported sex(31), only including samples with XX or XY chromosome pairs.

**Race:** Self-reported ethnic background(21000).

**Systolic Blood Pressure:** Systolic blood pressure, automated reading(4080)

**Diastolic blood pressure:** Diastolic blood pressure, automated reading(4079)

**Total Cholesterol:** Cholesterol(30690), converted to mg/dL.

**HDL Cholesterol:** HDL cholesterol(30760), converted to mg/dL.

**LDL Cholesterol:** LDL direct(30780), converted to mg/dL.

**History of Diabetes:** Using the combination of the (general) diabetes definition in section S4.1, only assigning case if onset was prior to assessment center visit, and the self-reported use of insulin(6177 coding 3).

**Smoker:** Smoking status(20116).

**Time since stopped smoking:** Time difference between the age field above and age stopped smoking(2897).

**On Hypertension Treatment:** Self-reported use of blood pressure medication(6177 coding 2) or any of the following medications listed in treatment/medication code(20003): chlorthalidone, chlorothiazide, hydrochlorothiazide, indapamide, metolazone, amiloride, spironolactone, triamterene, bumetanide, furosemide, torsemide, amiloride hydrochloride/hydrochlorothiazide, spironolactone/hydrochlorothiazide, triamterene/hydrochlorothiazide, acebutolol, atenolol, betaxolol, bisoprolol, bisoprolol/hydrochlorothiazide, metoprolol tartrate, metoprolol succinate, nadolol, pindolol, propranolol, solotol, timolol, benazepril, captopril, enalapril, fosinopril, lisinopril, moexipril, perindopril, quinapril, ramipril, trandolapril, candesartan, eprosartan, irbesartan, losartan, telmisartan, valsartan, amlodipine, diltiazem, felodipine, isradipine, nicardipine, nifedipine, nisoldipine, verapamil, doxazosin, prazosin, terazosin, carvedilol, labetalol, methyldopa, clonidine, guanfacine, hydralazine, minoxidil, eplerenone, spironolactone, and aliskiren.

**On a Statin:** Self-reported use of cholesterol lowering medication(6177 coding 1) or any medication containing "statin" in its name listed in treatment/medication code(20003).

**On Aspirin Treatment:** Any medication containing "aspirin" in its name listed in treatment/medication code(20003).

##### S4.3 Risk conversion for ASCVD predictors

The risk score output from the linear regression was converted into a risk estimate in percentage by the following steps.

1. The risk scores were calculated for the 185,106 samples used in training and binned into .05% quantiles. The actual disease prevalence was then calculated in each bin.
2. Two maps from risk scores to risk estimates were then calculated by pairing each risk score with its corresponding bin prevalence, sorting all these pairs according to risk score and then calculating the rolling average of the prevalence values using a triangular window. The two maps used window size 2000 and 9000, respectively. The map with the larger window was applied up to prevalence 0.53; above that limit the smaller window was required to capture the extremes.
3. The last step smoothed the conversion function by linear interpolation between each sample among the 100 lowest risk scores, then between each 2000th sample up to the $\sim 20,000$ highest risk scores where a step size of 2 was used.

##### S4.4 Coefficient sizes

The coefficient sizes for each (risk score | biomarkers) predictor can be found in figures **Figure S12** and **Figure S13**.

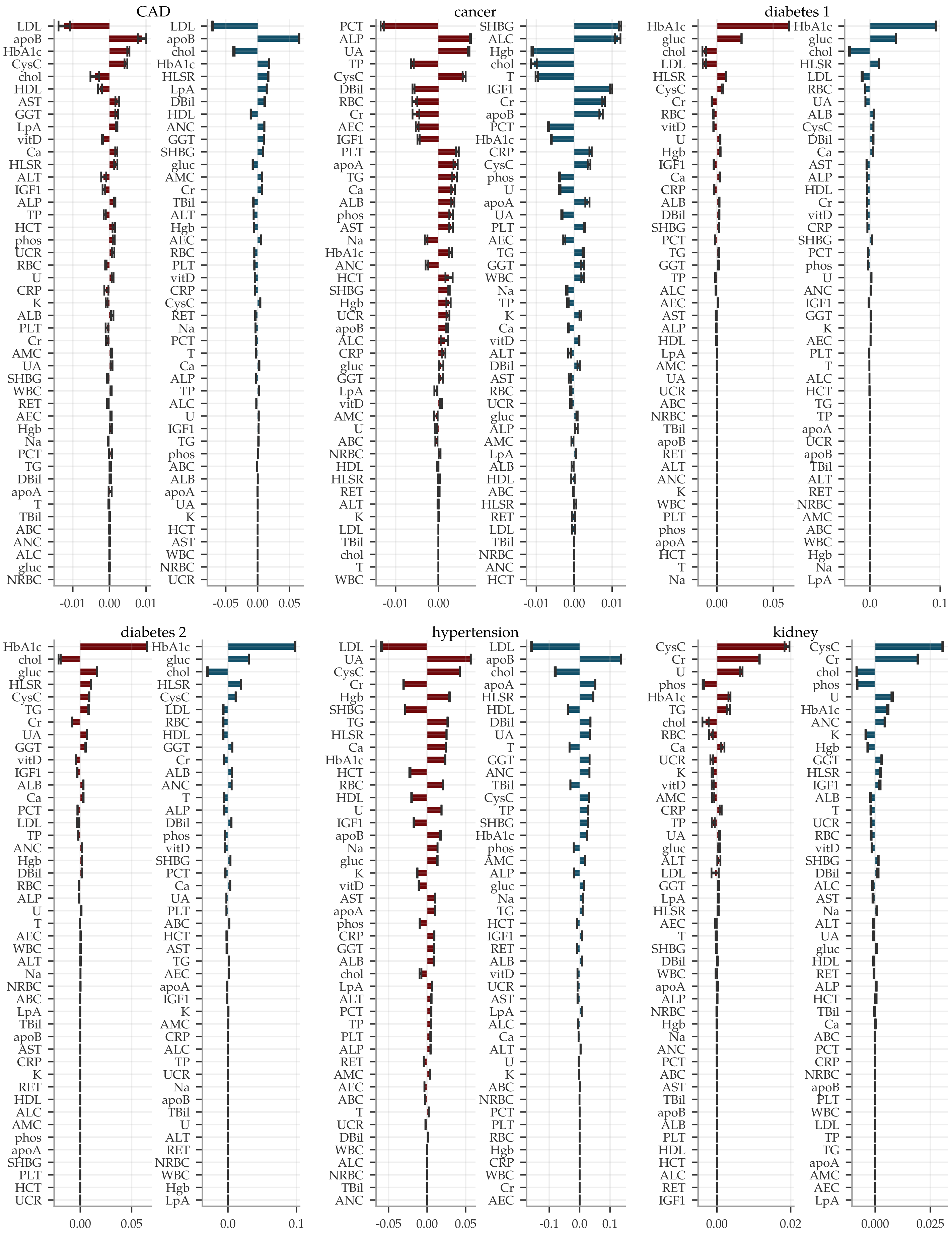


**Figure S12: Coefficient sizes (1 of 2) for (risk score | biomarkers) predictors, ■ women ■ men.** The bars show means of 5 predictors with error bars indicating the standard deviations. See **Figure S13** for the other two conditions.

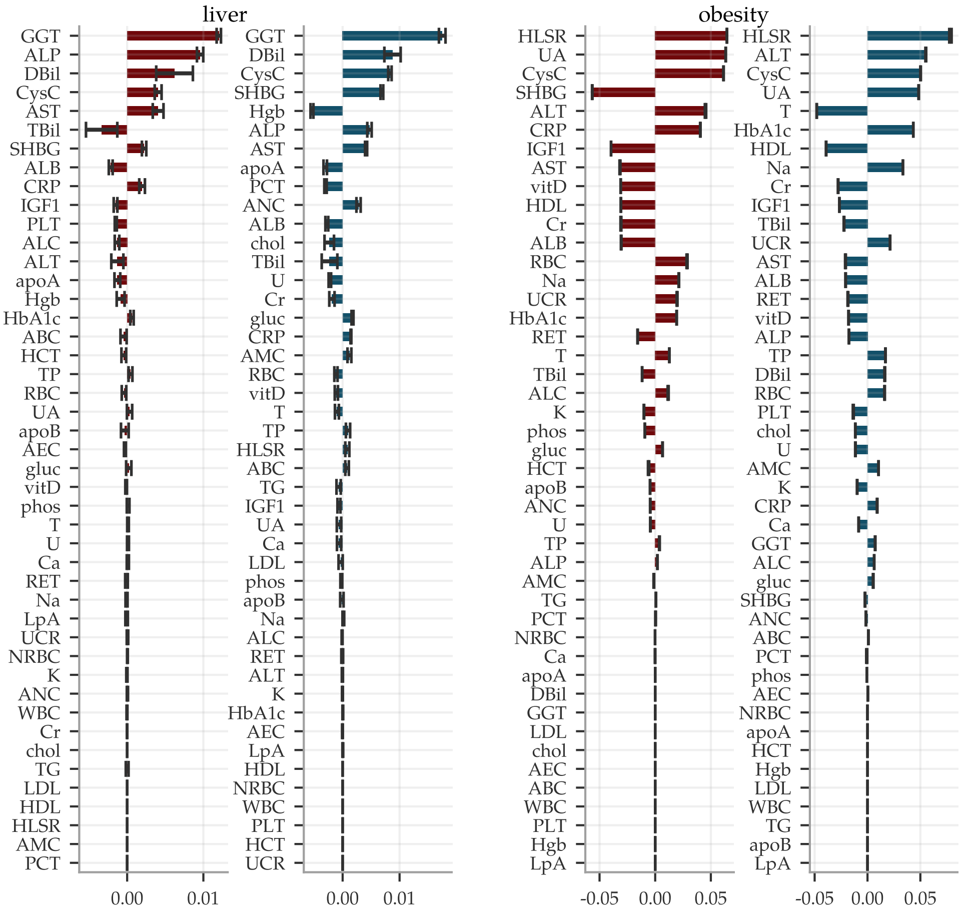


**Figure S13: Coefficient sizes (2 of 2) for the (risk score | biomarkers) predictors, ■ women ■ men.** The bars show means of 5 predictors with error bars indicating the standard deviations. The first six conditions are shown in **Figure S12**.

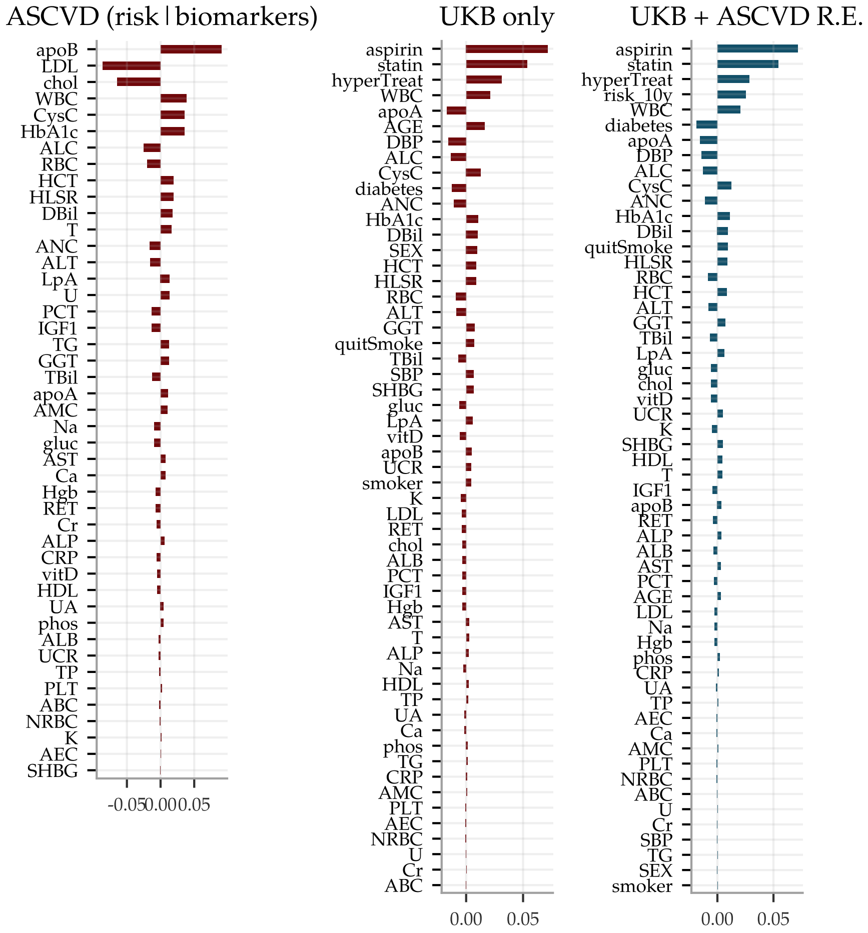


**Figure S14: Coefficient sizes for the different ASCVD predictors.** From left to right, (risk score | biomarkers) predictor for ASCVD, risk predictor using UKB data only but both biomarkers and all input used in the ASCVD Risk Estimator, and to the right using the biomarkers and both input and output for the ASCVD Risk Estimator.

##### S4.5 AUCs and sample sizes

The AUCs and sample sizes for the (risk score | biomarkers) predictors are shown in **Table S4**, while the sample sizes and numerical values for the AUCs of the (risk score | biomarkers | SNPs) predictors are listed in **Table S5**.

Sample sizes

| | women | | | | | | | | men | | | | | | | |
|---|---|---|---|---|---|---|---|---|---|---|---|---|---|---|---|---|
| | European | | South Asian | | East Asian | | African | | European | | South Asian | | East Asian | | African | |
| | ctrls | cases | ctrls | cases | ctrls | cases | ctrls | cases | ctrls | cases | ctrls | cases | ctrls | cases | ctrls | cases |
| CAD | 8842 | 181 | 1369 | 30 | 443 | 9 | 1361 | 18 | 8746 | 861 | 2329 | 365 | 329 | 9 | 1497 | 47 |
| cancer | 8076 | 940 | 1352 | 47 | 433 | 19 | 1323 | 56 | 8898 | 709 | 2642 | 52 | 329 | 9 | 1486 | 58 |
| diabetes 1 | 8785 | 232 | 1267 | 132 | 434 | 18 | 1280 | 99 | 9032 | 575 | 2212 | 482 | 320 | 18 | 1382 | 162 |
| diabetes 2 | 8702 | 314 | 1223 | 176 | 425 | 27 | 1239 | 140 | 8908 | 699 | 2148 | 546 | 319 | 19 | 1373 | 171 |
| hypertension | 6432 | 2584 | 968 | 431 | 339 | 113 | 751 | 628 | 5859 | 3748 | 1553 | 1141 | 242 | 96 | 906 | 638 |
| kidney | 8745 | 272 | 1363 | 36 | 441 | 11 | 1338 | 41 | 9112 | 495 | 2538 | 156 | 327 | 11 | 1463 | 81 |
| liver | 8754 | 264 | 1367 | 32 | 437 | 15 | 1347 | 32 | 9316 | 292 | 2607 | 87 | 321 | 17 | 1487 | 57 |
| obesity | 7121 | 1895 | 1073 | 326 | 434 | 18 | 763 | 616 | 7368 | 2239 | 2218 | 476 | 320 | 18 | 1077 | 467 |

AUCs

| | women | | | | men | | | |
|---|---|---|---|---|---|---|---|---|
| | European | South Asian | East Asian | African | European | South Asian | East Asian | African |
| CAD | 0.742 ± 0.002 | 0.756 ± 0.005 | 0.817 ± 0.004 | 0.688 ± 0.009 | 0.730 ± 0.000 | 0.743 ± 0.000 | 0.802 ± 0.002 | 0.692 ± 0.001 |
| cancer | 0.558 ± 0.001 | 0.659 ± 0.002 | 0.748 ± 0.006 | 0.660 ± 0.002 | 0.592 ± 0.001 | 0.628 ± 0.002 | 0.642 ± 0.003 | 0.634 ± 0.003 |
| diabetes 1 | 0.947 ± 0.000 | 0.936 ± 0.000 | 0.936 ± 0.001 | 0.918 ± 0.000 | 0.949 ± 0.000 | 0.931 ± 0.000 | 0.832 ± 0.001 | 0.909 ± 0.000 |
| diabetes 2 | 0.938 ± 0.000 | 0.917 ± 0.000 | 0.955 ± 0.000 | 0.891 ± 0.000 | 0.921 ± 0.000 | 0.880 ± 0.000 | 0.797 ± 0.001 | 0.886 ± 0.000 |
| hypertension | 0.710 ± 0.000 | 0.721 ± 0.000 | 0.766 ± 0.000 | 0.706 ± 0.000 | 0.716 ± 0.000 | 0.749 ± 0.000 | 0.670 ± 0.000 | 0.662 ± 0.000 |
| kidney | 0.697 ± 0.001 | 0.796 ± 0.003 | 0.740 ± 0.006 | 0.683 ± 0.002 | 0.666 ± 0.000 | 0.760 ± 0.000 | 0.791 ± 0.002 | 0.718 ± 0.001 |
| liver | 0.639 ± 0.003 | 0.691 ± 0.002 | 0.587 ± 0.011 | 0.647 ± 0.007 | 0.634 ± 0.001 | 0.689 ± 0.001 | 0.659 ± 0.002 | 0.688 ± 0.003 |
| obesity | 0.876 ± 0.000 | 0.821 ± 0.000 | 0.824 ± 0.000 | 0.811 ± 0.000 | 0.829 ± 0.000 | 0.749 ± 0.000 | 0.883 ± 0.000 | 0.747 ± 0.000 |

**Table S4: AUCs for (risk score | biomarkers) predictors.** Listed values are mean ± the standard deviation for 5 different predictors, all trained on European ancestry. These are the numerical values for figure 5 in the main document.

| | women | | | men | | |
|---|---|---|---|---|---|---|
| | AUC | ctrls | cases | AUC | ctrls | cases |
| CAD | 0.493 ± 0.010 | 2751 | 40 | 0.492 ± 0.001 | 2680 | 230 |
| cancer | 0.505 ± 0.002 | 2506 | 285 | 0.488 ± 0.005 | 2668 | 242 |
| diabetes 1 | 0.557 ± 0.003 | 2729 | 62 | 0.650 ± 0.002 | 2760 | 150 |
| diabetes 2 | 0.624 ± 0.009 | 2709 | 82 | 0.637 ± 0.003 | 2738 | 172 |
| hypertension | 0.558 ± 0.002 | 2074 | 717 | 0.514 ± 0.001 | 1831 | 1079 |
| kidney | 0.527 ± 0.005 | 2713 | 78 | 0.522 ± 0.003 | 2769 | 141 |
| liver | 0.524 ± 0.008 | 2719 | 72 | 0.551 ± 0.008 | 2830 | 80 |
| obesity | 0.574 ± 0.003 | 2236 | 555 | 0.564 ± 0.007 | 2309 | 601 |

**Table S5: AUCs and sample sizes for the (risk score | biomarkers | SNPs) predictors.** The AUCs are listed as means ± the standard deviations for 5 different predictors, trained and evaluated on European ancestry. These are the numerical values for figure 7 in the main document.

#### S4.6 Risk score distributions

The (risk score | biomarkers) and (risk score | biomarkers | SNPs) predictors were applied to evaluation sets of European ancestry with 9016/9607 and 2791/2910 women/men, respectively. The risk score distributions cases and controls for each sex are shown in **Figure S15** for (risk score | biomarkers) and **Figure S16** for (risk score | biomarkers | SNPs). Overlaid are QQ-plots for which cases and controls are separately compared to normal distributions of the same means and standard deviations to illustrate non-normally distributed deviations.

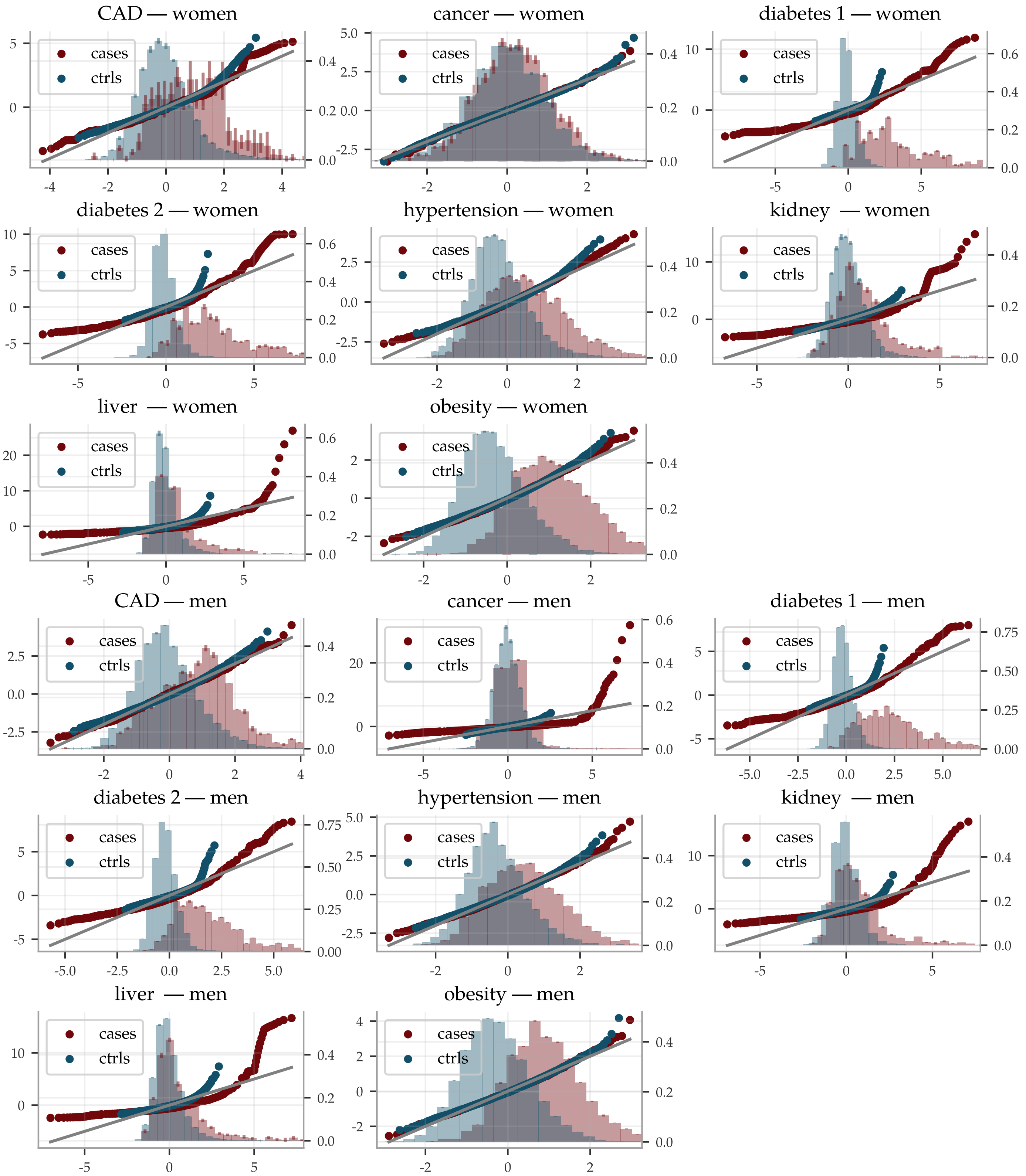


**Figure S15: QQ-plots and distribution histograms for the (risk score | biomarkers) predictors.** The QQ-plots compare case and control distributions with normal distributions of the same mean and standard deviations and use the y-axis to the left. The histograms are normalized to unit area and use the y-axis to the right, with error bars indicating the standard deviation for 5 different predictors. The evaluations are on 9016 women and 9607 men of European ancestry not used in training.

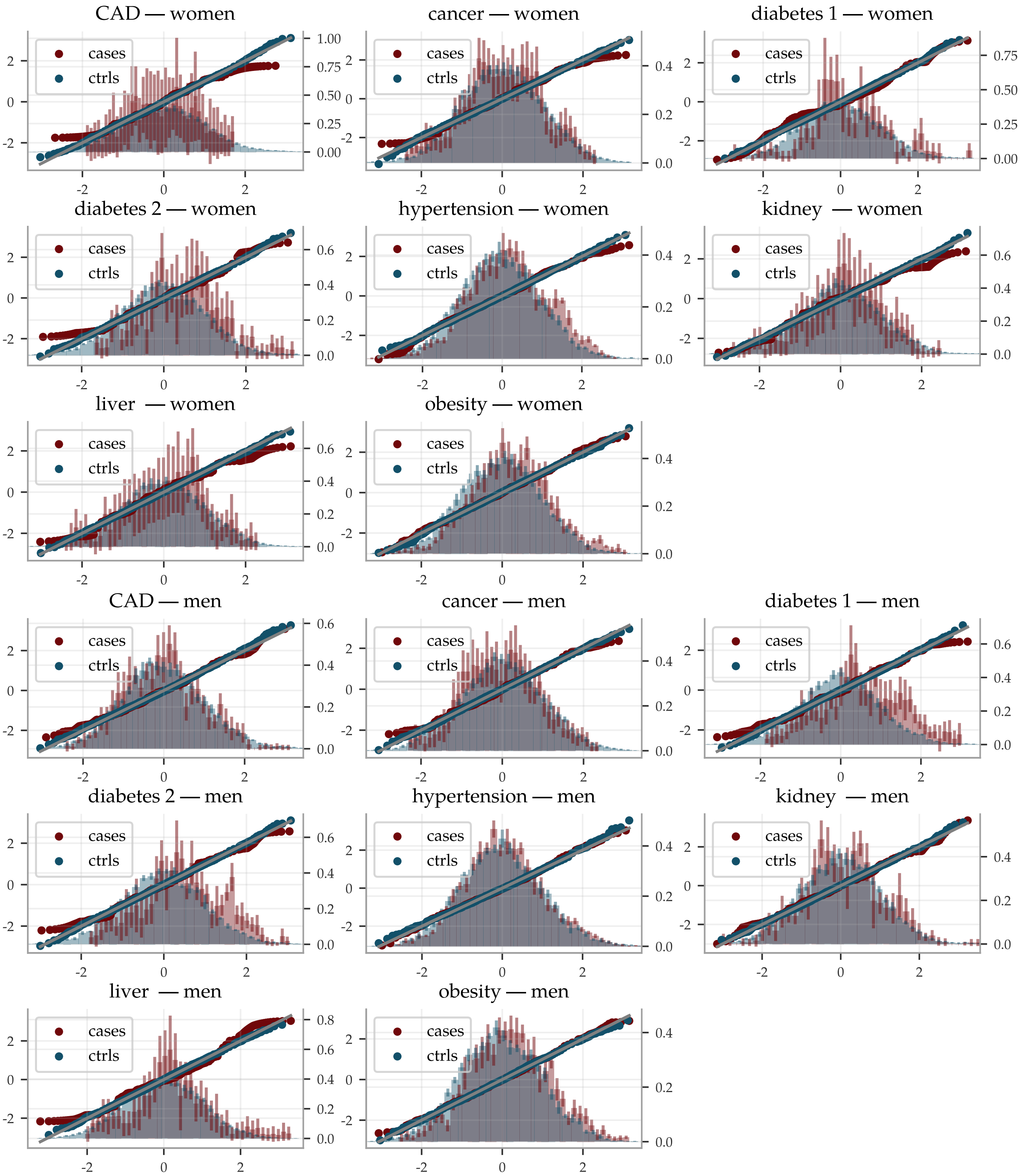


**Figure S16: QQ-plots and distribution histograms for the (risk score | biomarkers | SNPs) predictors.** The QQ-plots compare case and control distributions with normal distributions of the same mean and standard deviations and use the y-axis to the left. The histograms are normalized to unit area and use the y-axis to the right, with error bars indicating the standard deviation for 5 different predictors applied to 5 different PGS. The evaluations are on 2791 women and 2910 men of European ancestry not used in training.
